# Malaria and other risk factors for malnutrition, and the impact of intermittent preventive therapy for malaria on nutritional status of school-age children in Tanzania. A cross-sectional survey and a randomized controlled open-label trial

**DOI:** 10.1101/2024.01.03.24300756

**Authors:** Jeremiah John Hhera, Geofrey Makenga, Jean-Pierre Van geertruyden

## Abstract

**Background:** WHO and the Lancet reported that malaria and malnutrition form a double health burden in low and middle-income countries. Despite the massive implementation of several malaria interventions, there is limited information on the impact of intermittent preventive therapy (IPTsc) for malaria on the nutritional status of school-age children.

**Objective:** To determine malnutrition risk factors and evaluate the impact of IPTsc for malaria on the nutritional status of school-age children in North-East Tanzania.

**Methods:** We analyzed secondary data from a cross-sectional baseline survey and a randomized controlled open-label trial. Study participants were randomized to three treatment groups and thereafter followed for 20 months. Data were analyzed using logistic regression and a linear mixed model.

**Findings:** At baseline, the prevalence of malaria was 27%. 23% of ≤10 years children were underweight, 21% were stunted, and 28% were either thin or severely thin. The odds of stunting were 78% higher (AOR=1.78, 95%CI=[1.36, 2.33], P*<*0.001) among children who had malaria compared to those who did not. Children from low socioeconomic status (SES) had higher odds of being underweight (AOR=1.50, 95%CI=[1.13,2.01], P=0.006) compared to their high SES counterparts. During the intervention, change in mean weight, height, and BMI over time as estimated from age-treatment interaction was not significantly different in the DP and ASAQ treatment groups compared to the control group. A unit change in age increased weight, height, and BMI by 2.2 units (p-value *<*0.001), 3.3 units (p-value *<*0.001), and 0.5 units (p-value *<*0.001). The height and weight in female children were higher compared to that of male children by 1 unit (p-value *<*0.001) and 0.8 unit (p-value *<*0.001), respectively.

**Conclusion:** The burden of malaria and malnutrition in this study’s setting is remarkable. Instead of focusing only on malaria, public health agencies should reinforce nutritional programs by collaborating with local communities to ensure food availability in schools and provide sustainable nutritional education to the local community members.

## 1 Introduction

Malaria is a vector-transmitted disease predominantly distributed in tropical regions. It affects about 91 countries worldwide [1]. The World Health Organization (WHO) malaria report documented 227 million malaria cases in 2019 from 85 malaria-endemic countries and 241 million cases in 2020 [2, 3]. 96% of malaria cases and deaths were reported from the African region, and Tanzania was among the four countries that accounted for almost half the malaria-related deaths in 2020 [3]. A tremendous burden of malaria has been reported among children [2]. In malaria-endemic regions, a large proportion of school-age children live with subclinical malaria and thus a reservoir for malaria parasites [4]. In Muheza, Tanzania, children aged 5-14 years had a malaria prevalence of 39% in 2016 and 26.4% in 2019 [5, 6].

Children in lower and middle-income countries (LMICs) have been experiencing a double burden of malaria infections and malnutrition in the form of undernutrition. Malnutrition is a state of imbalance of intake of nutrients that leads to measurable adverse effects on body shape, functions, and clinical outcomes [7]. Body weight and height are the anthropometric indicators used for measurements to quantify nutritional status. To ensure consistency and comparability, the preferred units are the standard deviation (referred to as Z-score) of Weight-for-Age (WAZ), Height-for-Age (HAZ), and Body Mass Index (BMI) [8]. Growth and nutritional assessmentare primarily implemented worldwide in children of 0 to 5 years compared to older children and adolescents. UNICEF reported that 49.5 million children aged 0 to 5 years had acute malnutrition globally in2018 [9]. Although there is limited data on the nutritional status of 5 – 14 years old children, the Lancet report on worldwide body mass index (BMI) in 2016 documented the lowest record in South Asia and East Africa, 15.8 kg/m2 and 16.5 *kg/m*^2^, respectively [10]. The level of stunting is still high in LMICs, and the reduction rate is the lowest in the African region despite several interventional programs [9]. The level of stunting was reported to be high in 15 regions of the Tanzanian mainland in 2018 [11], and the lake zone prevalence of wasting was 11.3% and stunting was 29% [12].

Malaria-Malnutrition interaction is a complex phenomenon with an inconclusive causal relationship [13]. Malnutrition increases susceptibility to malaria, and recurrent malaria infections predispose the affected population to malnutrition [14]. Previous findings showed an increased risk of malaria in malnourished children. Malaria prevalence was higher among malnourished children of age ≤5 years compared to the ≥ 5 years group [15]. Observational studies from Sub-Saharan Africa and t h e Amazonian region reported an increased risk of malaria infection in undernourished school-age children [15, 16]. The relationship between stunting and malaria occurrence has been contradicting. As noted previously, stunting before transmission season was not associated with an increase in malaria incidence [17], and the other study reported that stunting is associated with reduced malaria incidence [16].

In the past two decades, malaria control programs have been massively implemented in disease-endemicareas [1, 2]. The use of drugs and vector-barrier for prevention was focused on ≤5 years and pregnant mothers [18], rarely involving school-age children. The previous interventions in both areas of intense perennial and seasonal transmission successfully minimized anemia and parasitemia in all age groups but did not examine the impact on nutritional status [4, 5, 19, 20]. However, it was noted in an observational study that treatment of severe malaria using artemisinin-based combinations improved nutritional status among children with severe acute malnutrition [21].

Generally, previous studies conducted mainly on children under five suggest an association between malaria and malnutrition. However, to the best of our knowledge, only a limited number of malaria-malnutrition studies included school-age children, and most malaria interventions did not address its impact on the nutritional status of school-age children. Therefore, we analyzed data from a baseline survey and randomized clinical trial using dihydroartemisinin-piperaquine (DP) and artesunate-amodiaquine (ASAQ) to, primarily, determine the association between malaria and malnutrition.

The hypothesis for this study was that there is an association between malaria and malnutrition in school-age children, and that the use of intermittent preventive therapy (IPTsc) for malaria improves nutritional status among school-age children. The specific objectives that guided our work were (i). To determine the prevalence of malnutrition and factors associated with malnutrition among school-age children (ii).To explore the attribution of malaria on low Weight-for-Age (WAZ), Height-for-Age (HAZ), and BMI in school-age children (iii). To evaluate the impact of IPTsc on the trend of weight, height, and BMI in school-age children.

## 2 Methods

### 2.1 Design and study population

We analyzed the secondary data from a cross-sectional baseline survey and a randomized controlled open-label trial. The preliminary study aimed to assess the effectiveness and safety of two anti-malarial drugs for malaria prevention in school children. In this 3-arm trial using a ”balanced block design,” Dihydroartemisinin-piperaquine (DP) and Artesunate-amodiaquine (ASAQ) were used with the ”standard of care” as a reference arm. The trial design and methods were described in a published study protocol [22] that was registered on clinical trials.gov-NCT03640403. The intervention continued for 12 months following recruitment. The study participants were followed for an extra 8 months to observe rebound effects and late adverse events due to interventional drugs.

The study targeted children of age between 5 to 15 years. Most children in this age range are enrolled in primary schools, making school-age children our source population. The following eligibility criteria were adopted from the study protocol [22], and children who met the criteria were included in the study.

#### 2.1.1 Inclusion criteria

- Male and female primary school children in a selected school
- Includes parental/guardian informed consent.
- Assent by primary school children aged 11 years and above.
- Aged 5–15 years at the baseline assessment.
- Currently, he/she lives within the pre-defined catchment area of Muheza District.
- Will remain within the same area throughout the study period (preferably pupils of class five and below).

#### 2.1.2 Exclusion criteria

Participants with at least one of the following criteria were excluded:

- Pupils of class/grade 6 or 7
- Currently enrolled in another study or participated in another investigational drug study within the last 30 days.
- Known to have heart disease or a known cardiac ailment.
- Reports known hypersensitivity to the study drugs or any sulphonamides.
- Not willing to undergo all study procedures including physical examination and to provide blood samples as per this study protocol.
- Having clinical features of severe anemia
- Has apparent severe infection or any condition that requires hospitalization
- Illness or conditions like haematologic, cardiac, renal, and hepatic diseases which in the judgement of the investigator would subject the participant to undue risk or interfere with the results of the study, including known G6PD deficiency and sickle cell traits.
- Body weight *<*14 kg

### 2.2 Study setting

The study was conducted in Muheza district, Tanga, Tanzania. The geographical location is in the northeastern part of Tanzania, where malaria transmission occurs almost throughout the year, full description can be accessed elsewhere [22].

### 2.3 Ethical approval

The study acquired regulatory approval from the Medical Research Coordination Committee (MRCC) of the National Institute of Medical Research (NIMR), Tanzania. The initial approval based on the protocol was granted on 20^th^ July 2018. The study protocol was amended and the committee reviewed and approved it on 19^th^ March 2019 with validity date on 19^th^ July 2019. However, based on the progress report of July 2019, the ethical clearance committee granted study extension until 20^th^ July 2020. The study approval numbers were NIMR/HQ/R.8a/Vol.IX/2818 and NIMR/HQ/R.8c/Vol.I/668 (for amendment) also NIMR/HQ/R.8c/Vol.I/1276 for ethical clearance extension.

### 2.4 Study procedures

During the baseline phase, pupils’ age, sex, level of education (grade in school), and residential address were recorded in case report forms after consent was granted. Given that participants were children, written informed consent was obtained from children’ parents. Children aged 11 or older were asked to assent. Further participants’ information and household characteristics were inquired from the parents when the study team visited homes to capture satellite locations using the Global Positioning System (GPS) and socioeconomic evaluation.

#### 2.4.1 Intervention

During the intervention, the treatment was either dihydroartemisinin-piperaquine (DP) or Artesunate amodiaquine (ASAQ). These drugs have no plausible causal relation to the nutritional status except when there is malaria infection as an intermediate variable. The malaria prevention dosage was given for three days in a directly observed manner, that is, by a study nurse and a trained teacher who would provide the drugs on subsequent days. Another tablet was given and documented when a child vomited within 30 minutes after ingesting a pill. Tanzania’s national malaria treatment and diagnostic guideline [23] considers DP and ASAQ as the alternative to first-line Artemether Lumefantrine (ALU) in treating uncomplicated malaria. In the case of malaria-confirmed illness, all children were treated using ALU except on the trial drugs dispensing days when those on DP and ASAQ groups were kept on the same drugsto treat uncomplicated malaria. The national guideline was followed in managing severe malaria [23]. Details on manufacturers’ instructions for these drugs, reporting, and handling of severe adverse events (SAE), and the laboratory procedures of malaria parasite investigation were described elsewhere [22].

#### 2.4.2 Outcome

The body weight and height that were measured during the trial were used to quantify nutritional status. A standard stadiometer and a weighing scale were used for measurements. The tools were calibrated by a trained staff before and after use to minimize measurement errors. The weight-for-age, height-for-age, and body mass index (BMI)-for-age Z-scores (WAZ and HAZ, ZBMI respectively) were calculated and used to classify the nutritional status. The BMI was first calculated as;

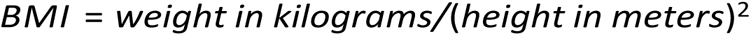

and the respective anthropometric index Z-scores were calculated using the ‘egen’ STATA function as described elsewhere [24, 25]. These Z-scores are the international standard for growth measurements derived by comparing individual growth measurements versus growth data from the reference or normal population, adjusted for age and sex. The standard deviations are normally categorized based on clinical relevance; *<*-3SD (severely underweight, severely stunted, severely thin), *<*-2SD to -3SD (underweight, stunted, thin), -2SD to +2SD (normal nutritional status), *>*+2SD to +3SD (overweight), and *>*+3SD (Obese) [26]. According to the WHO, WAZ nutritional index can be used up to the age of 10 years [26].HAZ can be used from birth through the adolescent period, and ZBMI Z-score is not recommended for the age of 2 years and below. The exposure-outcome relationship has been summarized in Figure 1 using Directed-Acyclic Graphs (DAGs).

**Figure 1:**
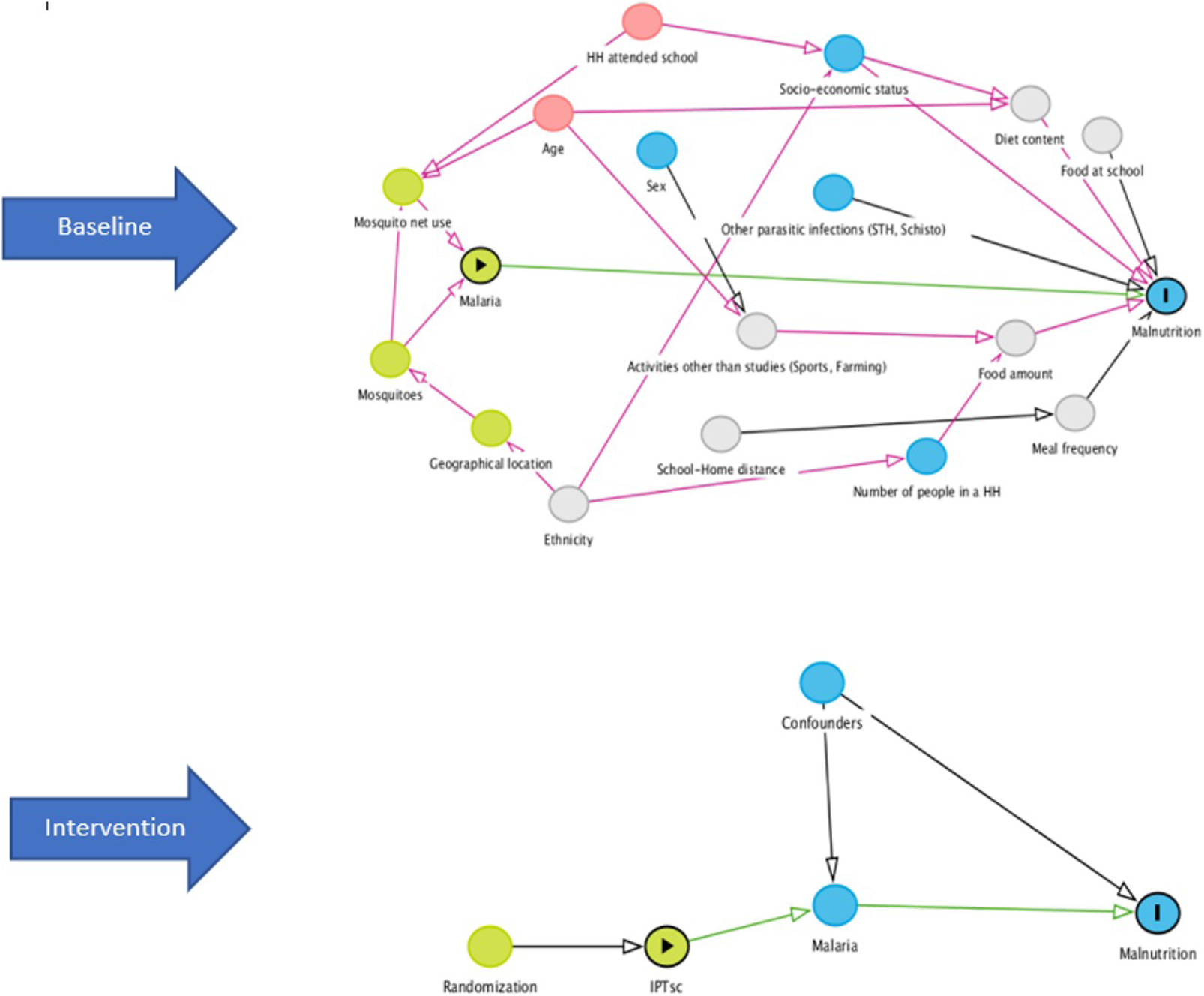
The Directed-Acyclic Graphs (DAGs) showing the exposure-outcome relationship for both baseline and interventional phase of our study.

### 2.5 Statistical Analysis

In this study, we analyzed baseline and longitudinal data that were collected from the same study participants. The methods of analysis were selected based on the nature of the data and the outcome variables ofinterest. Weight and height were summarized as median and categorical data were reported in proportions. Logistic regression and linear mixed models were applied in the data analysis to determine associations and the impact of anti-malaria prophylaxis on the nutritional status of children.

#### 2.5.1 Logistic regression

Both univariable and multi-variable logistic regressions were used to determine the association between malaria and other risk factorswith the nutritional indices. Univariable logistic regression was fitted to investigate the effect of a single covariate on causing underweight, stunting, or thinness status. For inclusion in the multivariable logistic model, covariates that had a p-value *<*0.25 were included as proposed by Hosmer and Lemeshow [27]. Multi-collinearity was checked using a generalized inflation factor (GVIF), and backward logistic regression was performed to build a parsimonious model. GVIF values were approximately 1 for all included variables, suggestive of no collinearity. In the process, a variable with a p-value greater than the significance level of 5% was dropped, and models were compared using a likelihood ratio test at each stage. A goodness of fit was checked using the Hosmer-Lemeshow goodness of fit test.

In addition, the attributable fraction (AF) (also known as attributable risk proportion (ARP)) was estimated for the variables that remained statistically significant in the adjusted analysis. By definition, AF is a measure that quantifies the proportion of the disease burden among exposed people that is caused by the exposure. We estimated AF from the logistic regression models as suggested by Greenland and Drescher [28, 29]. In the analysis of baseline data where malaria was an exposure of interest, AF was the proportion of prevalent underweight, stunting, or thinness that could be attributed to the malaria infection.

#### 2.5.2 Linear mixed model

In the analysis of longitudinal data, the intention-to-treat approach was implemented to evaluate the impact of anti-malarial treatment on nutritional status. Since the weight, height, and BMI measurements were repeated over time, the observations within each individual were assumed to be correlated. Such correlation violates the independence assumption of the classic linear model. To incorporate correlation within individuals and variation within and between children and schools, the linear mixed model was used in the analysis. The child identification number “child ID” and the “school “ were added as random effects to account for within-child variability due to repeated measurements and between-school variability. The correlation between children who live in the same households and variation between households was alsointeresting to study. However, we could not match children to their respective households and this could not be studied. This model formula was summarized in the following equation, which applies to all outcome parameters (weight, height, and BMI).

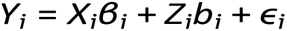

Where;

b_i_ ∼ N (0, D)

ɛ_i_ ∼ N (0, σ^2^)

b_1_, …b_n_, ɛ_i_, …ɛ_n_ are independent

Y_i_ = n_i_-dimensional vector for subject i, 1iN, where N is the number of subjects

X_i_ and Z_i_ are (n_i_p) and (n_i_q) dimensional matrices of known covariates.

β is the p-dimensional vector containing the fixed effects

b_i_ is the q-dimensional vector containing the random effects, and

ɛ_i_ is the dimensional vector containing residual components, and

D is the general (q × q) covariance matrix.

A study sample represents a population from which it was drawn; therefore, the distributional assumption was made for the random effects (*b_i_* ∼ *N* (0*, D*), *ɛ_i_* ∼ *N* (0*, σ*^2^)) with the covariance matrix D assumed to be unstructured. Variance components of random effects *b_i_* (diagonal elements of matrix D) measure the variability of longitudinal growth trajectories between individuals that are unexplained by covariates. The variability of repeated measurements within individual study participants was measured by the variance of residual component *ɛ_i_*.

In model building, we included the fixed effects of age, age as a quadratic term (age²), sex, the allocated treatment (study group), SES, and the interaction term of age-study group. The analysis model was decided by comparing the models with (in addition to fixed effects) random intercept and slope against models without random slope using the akaike information criterion (AIC). The model with random intercepts of ”child id” and ”school” had the lowest AIC value. The quadratic term of age was previouslyadded due to the difference in growth rate that causes a non-linear relationship between weight/height and age. The addition of quadratic term in the model introduced complexity in the interpretation. Its significance was further checked using AIC to compare a model with age and other covariates as described against the model with quadratic term included. The lower AIC was observed in the model that included a quadratic term of age. However, the difference was so small (see Appendix Table 12) that we ignored it and our final analysis model did not include a quadratic term. The estimates obtained from age-study group interaction were the effect sizes due to treatment over time. Assumptions for the linear model were checked by assessing the residual plots and mean of residuals, figures 8 and 9. The statistical inference employed in this analysis based on the restricted maximum likelihood method [30]. These analyses were carried out in R software version 4.2.1 for Windows, and the significance level was 0.05.

#### 2.5.3 Missing data management

Overall data missingness was about 5% during the study. The reason for fewer data missingness was that visits were done on regular schooling days at the respective schools. The missing pattern affected individuals from two schools where scheduled visits were canceled as the government ordered sudden school closure due to covid-19 outbreak. Public activities were resumed within three months and the study visits continued accordingly. The reason for missingness did not relate to observed characteristics or the missing data; hence, missing data completely at random (MCAR) was considered the missing data mechanism. Considering the low rate of missingness and MCAR data missing mechanism, complete case analysis was applied.

## 3 Results

### 3.1 General characteristics of the study population

In February 2019, 2628 children aged 0-15 years were recruited from 958 households for socioeconomic and demographic assessment. The study focused on children of age 5-15 years; therefore, primary school children were a target source population. 1566 school-age children who met inclusion criteria and signed a consent form were clinically evaluated during the baseline phase of the study. Among them,826 (53%) were male, and 740 (47%) were female children. The children’s age ranged from 5 to 15 years, with the median age being 9 years. The weight and height of all children were measured during the baseline data collection, and their median values were 23Kgs and 128.5cm, respectively. The mean BMI was 14.98(2.49)*kg/m*^2^. 1566 children were further assigned to three treatment groups of two anti-malarial drugs and a standard care or control group. The balanced three study groups were Dihydroartemisinin-Piperaquine (DP) (n=526), Artesunate-Amodiaquine (ASAQ) (n=527), and Standard Care or control group (n=513). The average weight gains during the study period for the DP, ASAQ, and Standard Care treatment groups were 5kg, 5.1kg, and 5.2kg, respectively. For the children’s height, the net gains were 6cm, 6.1cm, and 6.5cm in the DP, ASAQ, and Standard Care treatment groups, respectively.

### 3.2 Distribution of participants’ characteristics and their nutritional outcomes

The extreme categories of weight-for-age (severely underweight), height-for-age (severely stunted), BMI-for-age (severely thin), and obesity were in a very small (*<*5%) proportion in this study population. In this analysis, we used two categories, that is, WAZ *<*-2 (underweight) and WAZ ≥-2 (normal weight). 246 (23%) of 1064 children of age 5 to 10 years were either underweight or severely underweight. 327 (21%) of all study participants were stunted, and 439 (28%) were either thin or severely thin. Among 1566 children, 806 (51%) belong to low SES households, 1352 (86%) heads of household (HH) acquired at least primary education, and 216 (14%) did not attend school. Regarding infections, 403 (27%) of 1484 who were tested had malaria. 661 (42%) of all children reported a history of malaria in the past month, and 210 (13%) used anti-malarial drugs in the previous week. When screened for other parasites, the prevalence of soil-transmitted helminths (STH) and schistosomiasis were 1% and 8%, respectively. Table 1 provides the summary of variables and their distribution proportions.

**Table 1:**
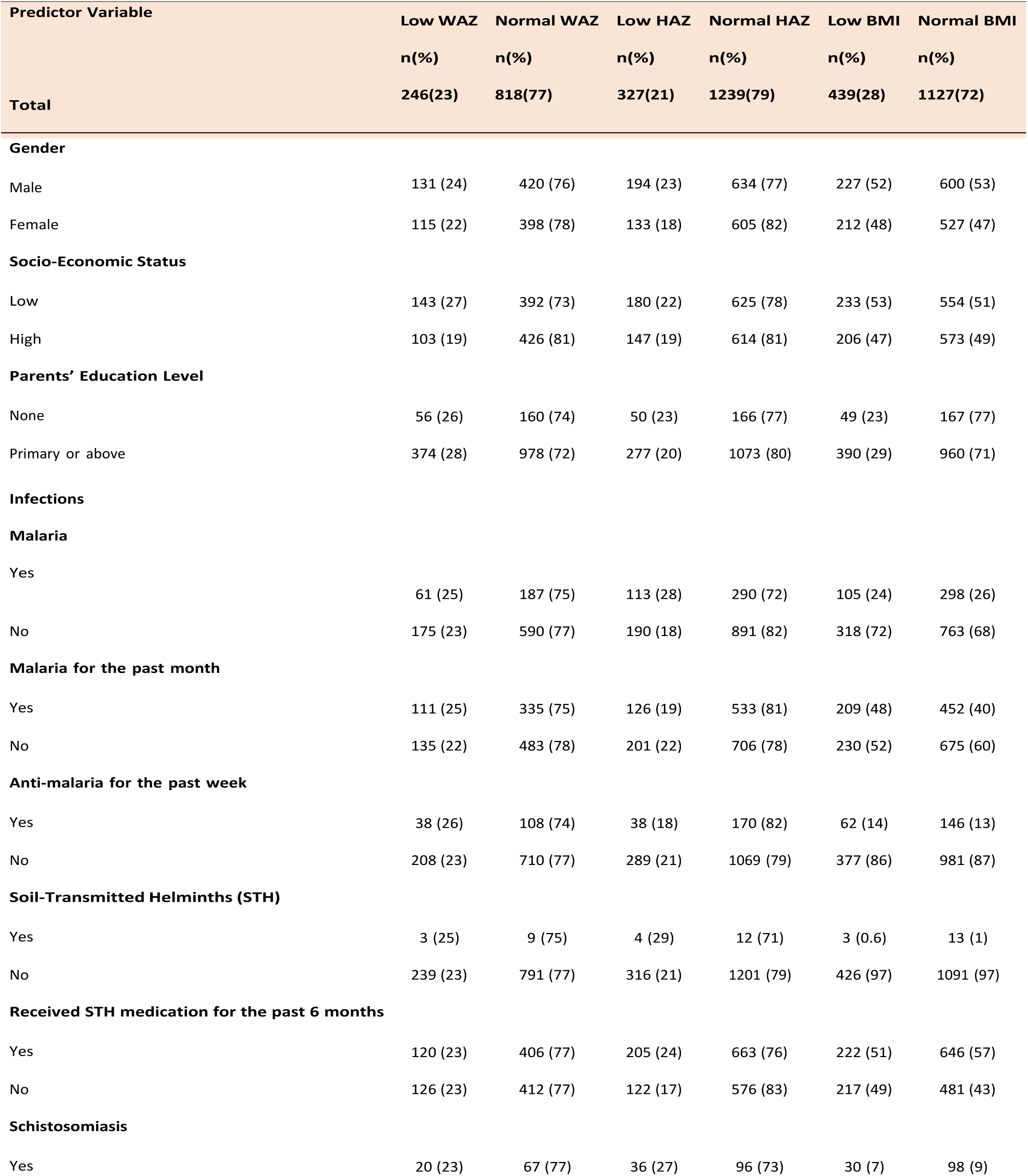
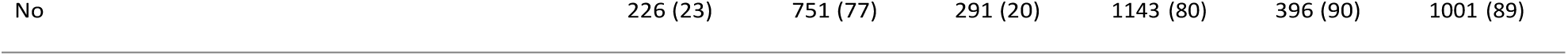
Baseline characteristics of study participants.

During the intervention, children’s weight and height were measured at all visits, and malaria testing was done at baseline, after the first 12 months, and at the 20th month of study. Due to the absence of data for malaria at several scheduled visits, we only compared the point prevalence of nutritional indices and malaria at baseline and after 12 months. The point prevalence of malaria and nutritionalstatus according to treatment allocation were summarized in Table 2 and Figure 2. Generally, malaria prevalence was notably low after 12 months of intervention in groups that received treatments compared to the standard care group. The underweight, stunted, and thin status among children did not largely vary from baseline to the 12th-month visit.

**Figure 2:**
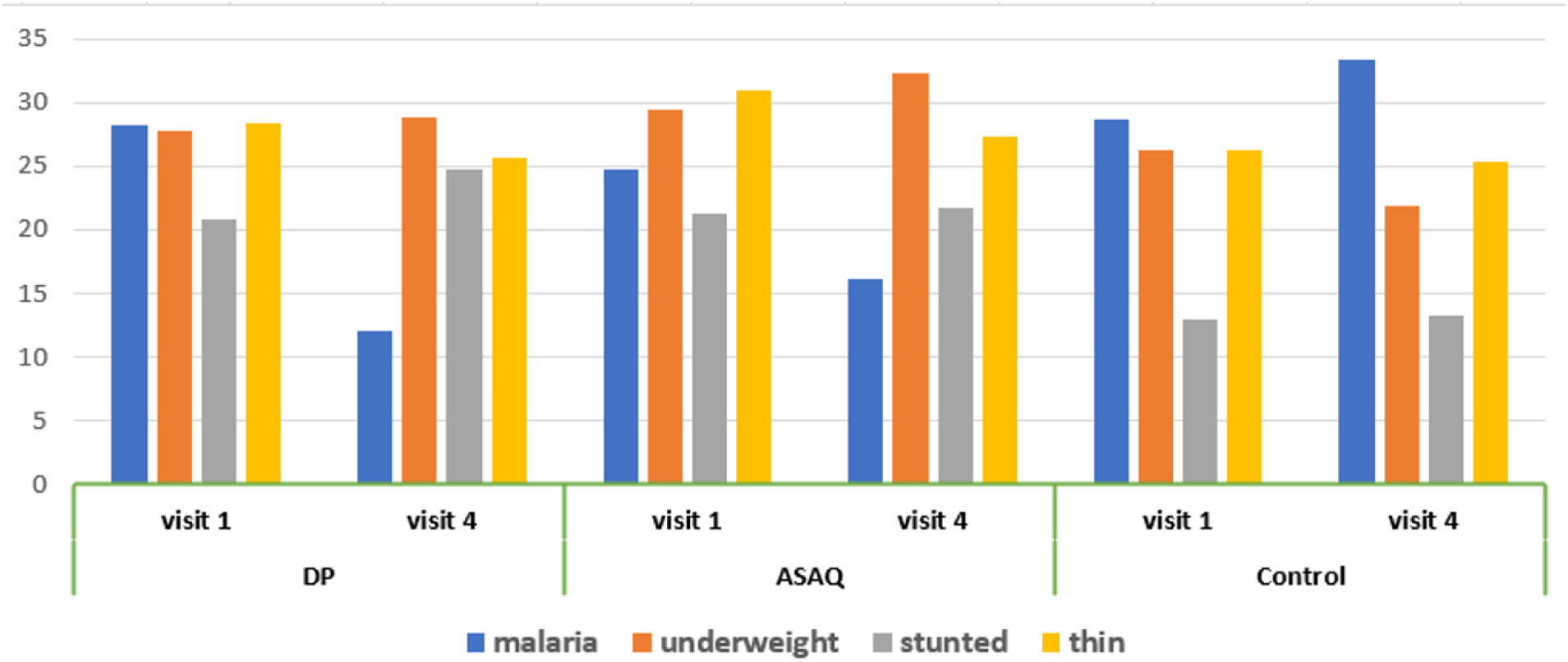
Trend of malaria and nutritional status before and after intervention

**Table 2:**
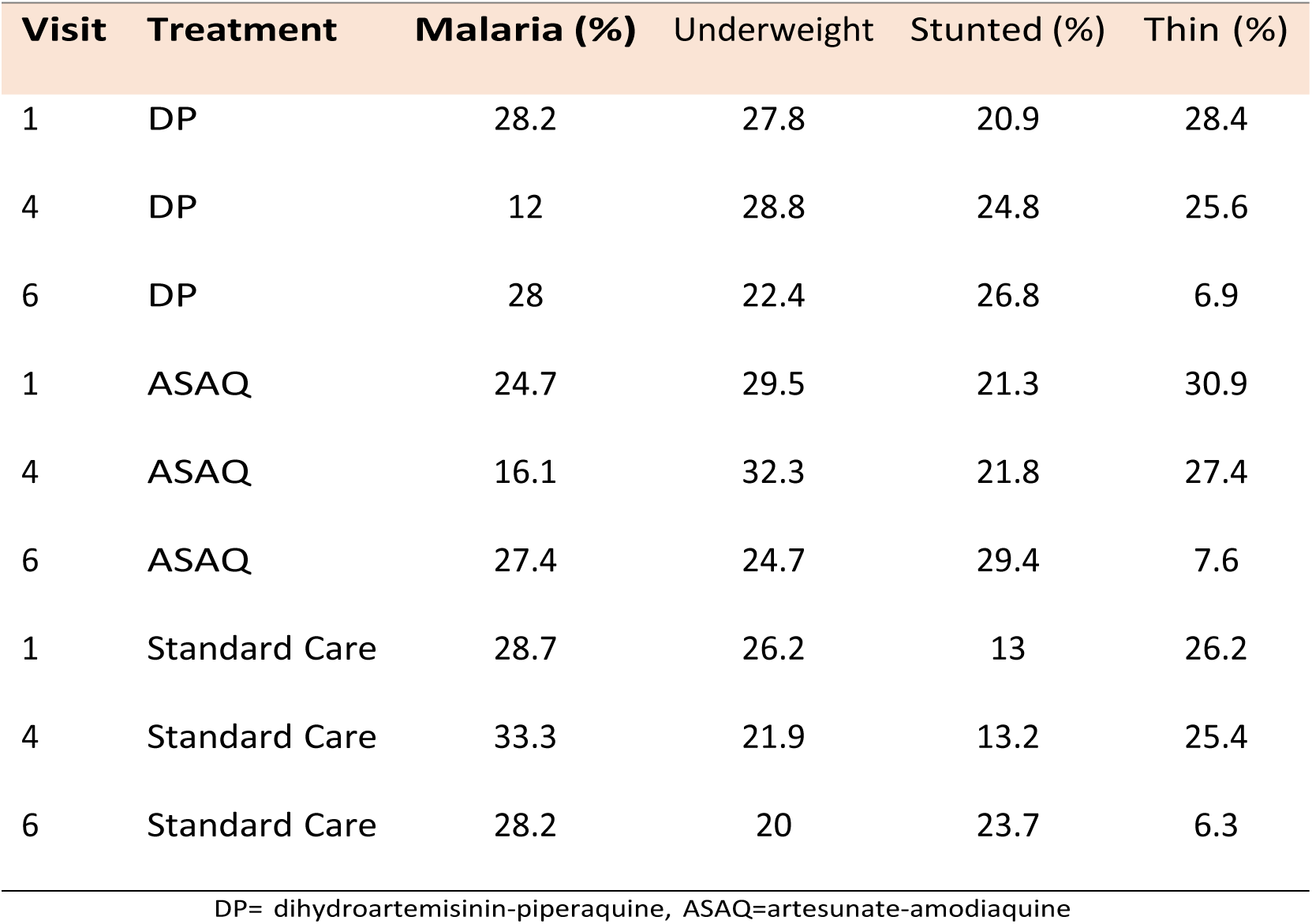
Trend of malaria infections between study groups during follow-up visits.

| Visit | Treatment | Malaria (%) | Underweight | Stunted (%) | Thin (%) |
| --- | --- | --- | --- | --- | --- |
| 1 | DP | 28.2 | 27.8 | 20.9 | 28.4 |
| 4 | DP | 12 | 28.8 | 24.8 | 25.6 |
| 6 | DP | 28 | 22.4 | 26.8 | 6.9 |
| 1 | ASAQ | 24.7 | 29.5 | 21.3 | 30.9 |
| 4 | ASAQ | 16.1 | 32.3 | 21.8 | 27.4 |
| 6 | ASAQ | 27.4 | 24.7 | 29.4 | 7.6 |
| 1 | Standard Care | 28.7 | 26.2 | 13 | 26.2 |
| 4 | Standard Care | 33.3 | 21.9 | 13.2 | 25.4 |
| 6 | Standard Care | 28.2 | 20 | 23.7 | 6.3 |

DP= dihydroartemisinin-piperaquine, ASAQ=artesunate-amodiaquine

**Table 3:** Risk factors of being underweight (WAZ*<*-2) in a malaria endemic region among school-age children.

| Variable<sup>a</sup> |  | Crude OR | 95% CI | p-value | Adjusted OR<sup>b</sup> | 95% CI | p-value | ARP |
| --- | --- | --- | --- | --- | --- | --- | --- | --- |
| Sex | Male | 1.07 | 0.81, 1.43 | 0.622 |  |  |  |  |
|  | Female | Ref |  |  |  |  |  |  |
| Socio-Economic Status | Low | 1.50 | 1.13, 2.01 | 0.005 | 1.50 | 1.13, 2.01 | 0.006* | 0.27 |
|  | High | Ref |  |  |  |  |  |  |
| Number of people in a household |  | 1.0 | 0.93, 1.07 | 0.988 |  |  |  |  |
| Head of HH attended school | Yes | 0.92 | 0.62, 1.40 | 0.699 |  |  |  |  |
|  | No |  |  |  |  |  |  |  |
| Malaria parasitemia | Yes | 0.99 | 0.76, 1.27 | 0.929 |  |  |  |  |
|  | No | Ref |  |  |  |  |  |  |
| Malaria in the past Month | Yes | 1.19 | 0.89, 1.59 | 0.235 | 1.20 | 0.90, 1.59 | 0.226 |  |
|  | No | Ref |  |  |  |  |  |  |
| Soil-Transmitted Helminths | Yes | 0.85 | 0.49, 1.34 | 0.497 |  |  |  |  |
|  | No | Ref |  |  |  |  |  |  |
| Schistosomiasis | Yes | 1.00 | 0.70, 1.39 | 0.990 |  |  |  |  |
|  | No | Ref |  |  |  |  |  |  |

OR=Odds Ratio, CI=Confidence Interval, HH=Head of Household, Ref= Reference category, \* significant p-value, ARP=
Attributable Risk Proportion. <sup>a</sup>Variables were selected based on the previous literature and pre-analysis Directed-Acyclic Graphs
(DAGs) Figure 1. <sup>b</sup>Results are adjusted OR from the multivariable logistic regression model.

### 3.3 Malaria and Other Risk Factors of Malnutrition in School-age Children

#### 3.3.1 Malaria and other risk factors of being underweight (WAZ*<* -2) in school-age children

Among children diagnosed with malaria parasites, 61 (25%) of 248 were either underweight or severely underweight (Table 1). Table 2 summarizes the results of risk factors of being underweight from both univariable and multivariable logistic regression analysis. In univariable logistic regression, children from low SES households had 1.5 times higher odds (95% CI= [1.13, 2.01], P=0.005) of being underweight than the high SES category. The association between underweight status and low SES remained statistically significant even in the adjusted analysis (95% CI= [1.13, 2.01], P=0.006).

#### 3.3.2 Malaria and other risk factors of stunting (HAZ *<* -2) in school-age children

113 (28%) of all children with malaria were stunted (Table 1). In univariable logistic regression (Table 4), the odds of being stunted in the male sex were 1.4 times (OR=1.40, 95% CI=[1.09, 1.79], P=0.008) higher compared to females. Children diagnosed with malaria had about 1.6 times higher odds of being stunted compared to those who did not have malaria (OR=1.55, 95% CI=[1.55, 1.88], P*<*0.001). When adjusted for “sex” in the multivariable logistic regression, children with malaria infection had 78% higher odds (OR=1.78, 95% CI=[1.36, 2.33], P*<*0.001) of stunting compared to those who did not have malaria. Malaria was attributed to stunting by 37% in this study population.

**Table 4:** Risk factors of stunting (HAZ*<*-2) in a malaria endemic region among school-age children.

| Variable<sup>a</sup> |  | Crude OR | 95% CI | p-value | Adjusted OR<sup>b</sup> | 95% CI | p-value | ARP |
| --- | --- | --- | --- | --- | --- | --- | --- | --- |
| Sex | Male | 1.40 | 1.09, 1.79 | 0.008 | 1.36 | 1.06, 1.74 | 0.017* | 0.23 |
|  | Female | Ref |  |  |  |  |  |  |
| Socio-Economic Status | Low | 1.20 | 0.94, 1.54 | 0.141 |  |  |  |  |
|  | High | Ref |  |  |  |  |  |  |
| Number of people in a household |  | 1.03 | 0.97, 1.09 | 0.289 |  |  |  |  |
| Head of HH attended school | Yes | 0.86 | 0.61, 1.21 | 0.385 |  |  |  |  |
|  | No | Ref |  |  |  |  |  |  |
| Malaria parasitemia | Yes | 1.55 | 1.28, 1.88 | <0.001 | 1.78 | 1.36, 2.33 | <0.001* | 0.37 |
|  | No | Ref |  |  |  |  |  |  |
| Malaria in the past Month | Yes | 0.75 | 0.40, 1.34 | 0.134 |  |  |  |  |
|  | No | Ref |  |  |  |  |  |  |
| Soil-Transmitted Helminths | Yes | 1.04 | 0.68, 1.51 | 0.853 |  |  |  |  |
|  | No | Ref |  |  |  |  |  |  |
| Schistosomiasis | Yes | 1.13 | 0.85, 1.49 | 0.396 |  |  |  |  |
|  | No | Ref |  |  |  |  |  |  |

#### 3.3.3 Malaria and other risk factors of being thin (ZBMI*<* -2) in school-age children

Among 403 children who had malaria, 105 (26%) were thin as they were in a category of less than negative 2 BMI Z-score (Table 1). The effect sizes for the association between thinness and the risk factors are provided in Table 5. The results of univariable logistic regression showed that the odds of being thin were 36% higher (OR=1.36, 95% CI=[1.09, 1.70], P*<*0.001) in children who reported a history of malaria illness in the past month. In the adjusted analysis, the association between history of malaria infection and thinness remained significant (AOR=1.37, 95% CI=[1.10, 1.71], P=0.006). The association between head of household education level and thinness in children was not significant in both univariable and multivariable logistic regression.

**Table 5:**
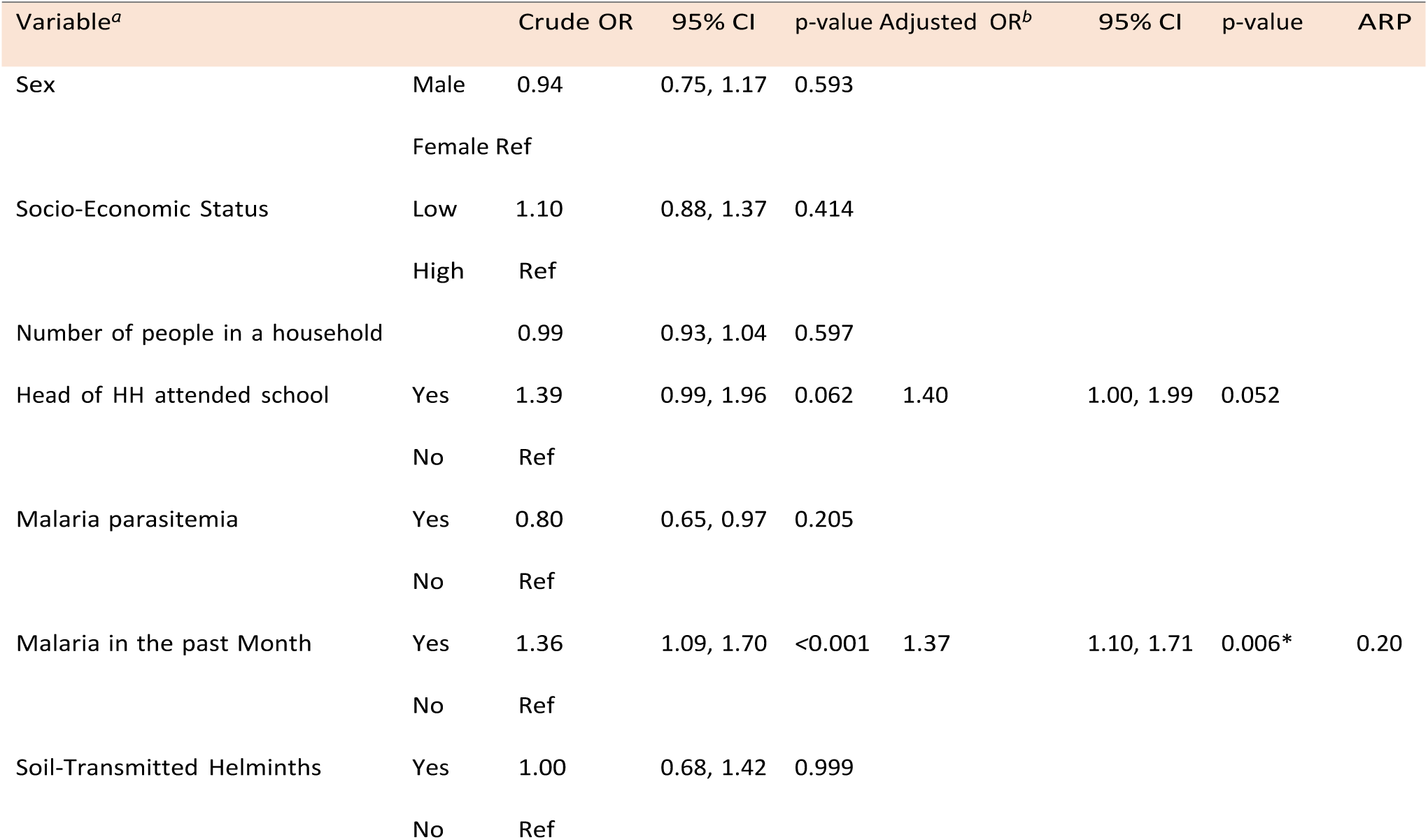
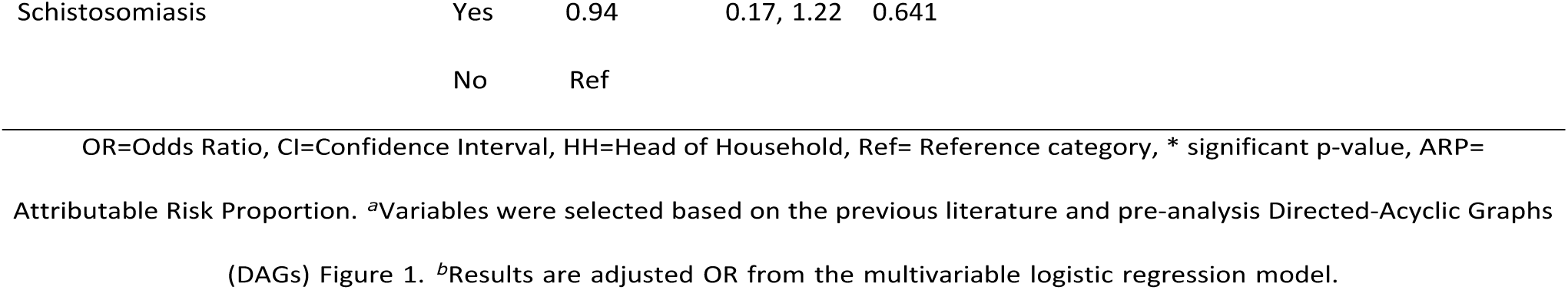
Risk factors of being thin (ZBMI*<* -2) in a malaria endemic region among school-age children.

| Variable<sup>a</sup> |  | Crude OR | 95% CI | p-value | Adjusted OR<sup>b</sup> | 95% CI | p-value | ARP |
| --- | --- | --- | --- | --- | --- | --- | --- | --- |
| Sex | Male | 0.94 | 0.75, 1.17 | 0.593 |  |  |  |  |
|  | Female | Ref |  |  |  |  |  |  |
| Socio-Economic Status | Low | 1.10 | 0.88, 1.37 | 0.414 |  |  |  |  |
|  | High | Ref |  |  |  |  |  |  |
| Number of people in a household |  | 0.99 | 0.93, 1.04 | 0.597 |  |  |  |  |
| Head of HH attended school | Yes | 1.39 | 0.99, 1.96 | 0.062 | 1.40 | 1.00, 1.99 | 0.052 |  |
|  | No | Ref |  |  |  |  |  |  |
| Malaria parasitemia | Yes | 0.80 | 0.65, 0.97 | 0.205 |  |  |  |  |
|  | No | Ref |  |  |  |  |  |  |
| Malaria in the past Month | Yes | 1.36 | 1.09, 1.70 | <0.001 | 1.37 | 1.10, 1.71 | 0.006* | 0.20 |
|  | No | Ref |  |  |  |  |  |  |
| Soil-Transmitted Helminths | Yes | 1.00 | 0.68, 1.42 | 0.999 |  |  |  |  |
|  | No | Ref |  |  |  |  |  |  |

|  |  |  |  |  |
| --- | --- | --- | --- | --- |
| Schistosomiasis | Yes | 0.94 | 0.17, 1.22 | 0.641 |
|  | No | Ref |  |  |

OR=Odds Ratio, CI=Confidence Interval, HH=Head of Household, Ref= Reference category, \* significant p-value, ARP=

### 3.4 The trend of weight, height, and body mass index (BMI) during and after the trial

#### 3.4.1 The average trend of children’s height, weight, and BMI

The children’s weight, height, and BMI were measured prior to starting the 12 months intervention. It was thereafter measured during each follow-up visit, including the eight months post-trial. Figure 3 shows the evolution of mean height predicted from the linear mixed model across age. Overall, the children’s height increased almost linearly with increasing age. Visually, there is no significant difference in the trend of average height for age between the study groups. Due to similarities between children randomized to different treatment groups, their mean height data points overlap, markedly in the 5 to 10 years age group. The plots for the weight (Figure 6), BMI (Figure 7), and the trend of height-for-age grouped by sex (Figure 5) are available in the appendix.

**Figure 3:**
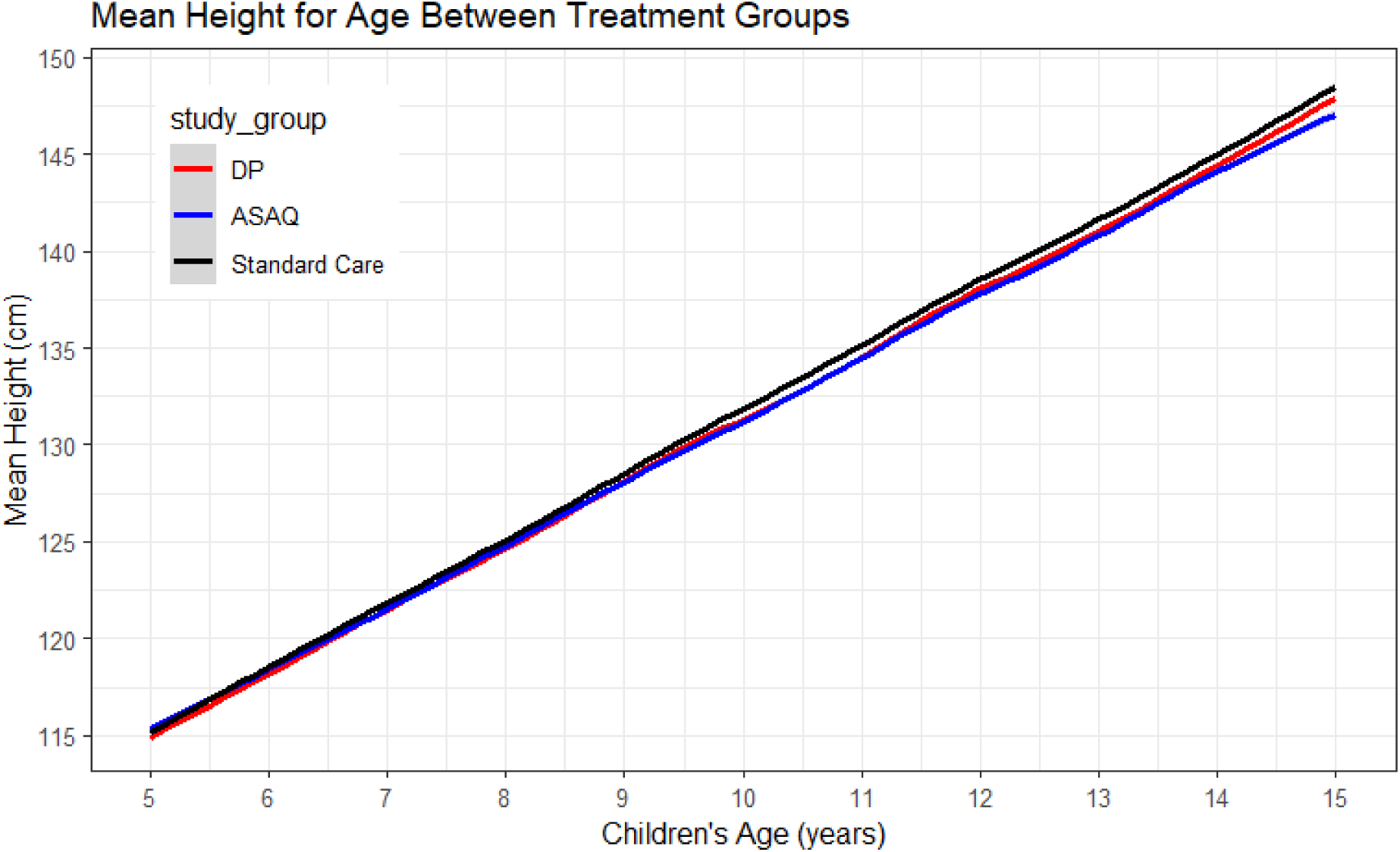
The trend of mean height-for-age of school-age children grouped by treatment

#### 3.4.2 The individual profile for the children’s height, weight, and BMI

Change in children’s height, weight, and BMI at the individual level was explored using profile plots. A random sample of 70 children was drawn and their individual plots were plotted in Figure 4. The plots showed minimum data imbalance as there was not much noticeable discontinuation of individual lines at subsequent data points, which signifies minimum missingness in the data. Following individual profile lines, the data points of many subjects inconsistently change from lower to higher values and vice versa. Such a pattern of changes signifies high within-child variability among individuals as their height, weight, andBMI evolved with time. High variability between children was also seen in plots a, b, and c.

**Figure 4:**
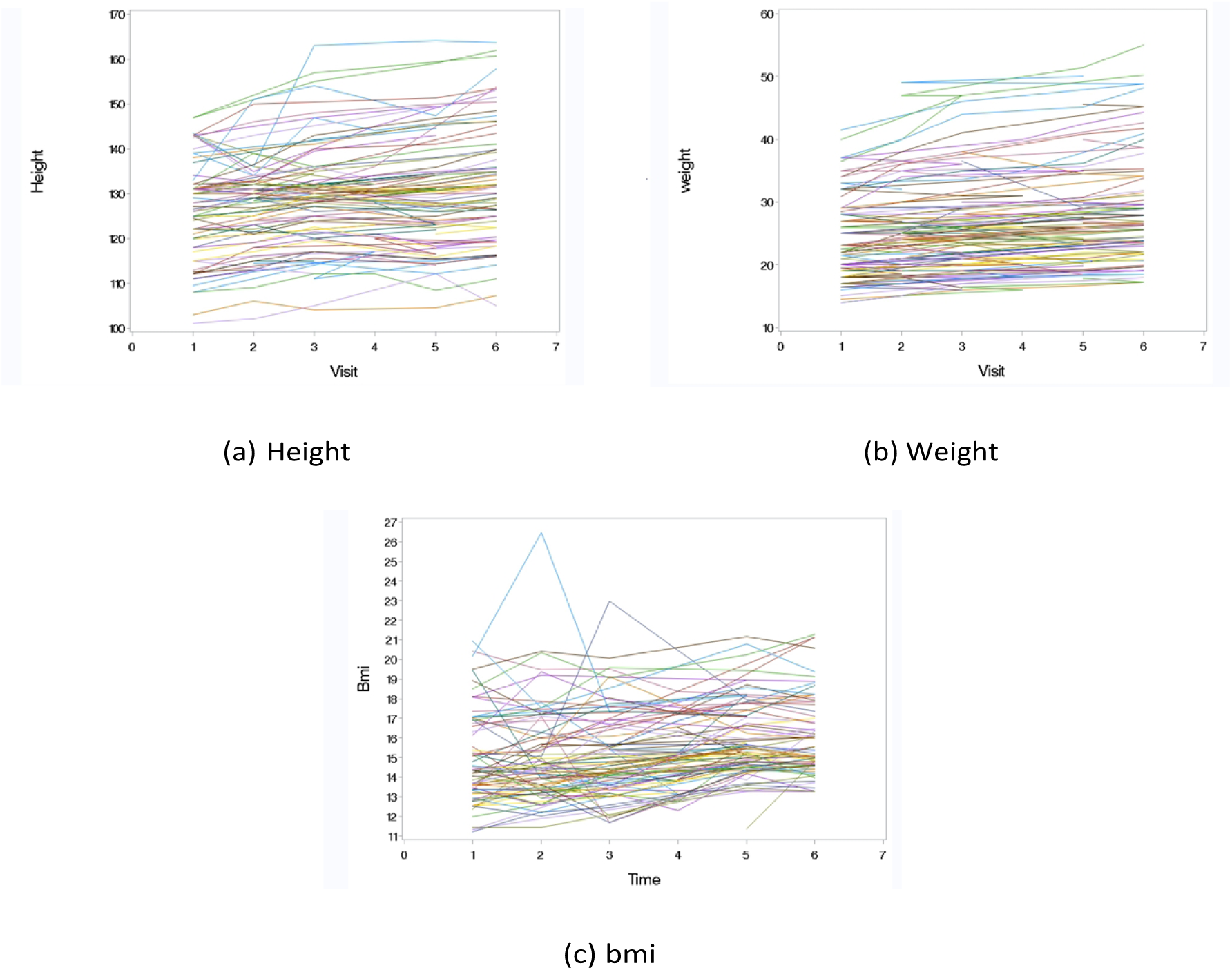
Individual profile plots for 70 randomly selected children

#### 3.4.3 The impact of malaria intermittent preventive therapy on weight, height, and BMI

The impact of malaria preventive treatment on growth indices was investigated using the linear mixed model (LMM). Different models with and without random effects were compared using AIC and the results are presented in Appendix Table 13. The model with a “child ID” and “school” as random intercept had the smallest AIC, and was therefore selected as the final model.

Our analysis that investigated random effects of individual ”child id” and ”school” revealed that between-children and within-child variability were high, whereas between-schools variability was minimal for all growth indices. Quantitatively, the between-children (child id) variance (Tables 6, 7, and 8) were 18.06 (81%), 25.95 (61.4%), and 9.82 (26.4%) for weight, height, and BMI, respectively. The between-schools variance was less than 1% for weight, height, and BMI. Repeated measurements for each study participant also explained a large portion of variance as estimated by the residual component of the LMM. The within-child variance in weight, height, and BMI were 4.1 (18.4%), 16.07 (38%), and 27.2 (73.2%), respectively. The proportions described here are relevant to the individual profile plots described in the previous section 3.4.2. The effect of malaria preventive therapy (tables 6, 7, and 8) on weight, height, and BMI was evaluated by including the treatment allocation as a fixed effect covariate (that provided the estimates for comparison at baseline), and the interaction term of treatment allocation and age (that provided estimates of changing slope over time). At baseline, the treatment was not consumed by the participants; therefore, the latter reflects the true treatment effect that was studied. Adjusted for other covariates in the model, the means of weight, height, and BMI in DP and ASAQ treatment groups were not significantly different from that of the control group at baseline. During the intervention, change in mean weight, height, and BMI over time as estimated from age-study group interaction was not significantly different in DP and ASAQ groups compared to the control group. A unit change in age increased weight, height, and BMI by 2.2 units (p-value *<*0.001), 3.3 units (p-value *<*0.001), and 0.5 units (p-value *<*0.001). The height and weight in female children were higher compared to that of male children by 1 unit (p-value *<*0.001) and 0.8 unit (p-value *<*0.001), respectively. Compared to males, being female did not significantly affect the BMI in our study. The change in weight, height, and BMI was not significantly different in children from low SES households compared to the high SES category.

**Table 6:** LMM results for weight during the trial.

| Random effects |  | Variance |  |  |  |
| --- | --- | --- | --- | --- | --- |
| Child id (intercept) |  | 18.0647 |  |  |  |
| School (intercept) |  | 0.1230 |  |  |  |
| Residual |  | 4.1008 |  |  |  |
| Fixed effect | estimate | sd | df | t value | p value |
| Intercept | 5.4418 | 0.7495 | 945 | 7.26 | <.0001* |
| age | 2.1758 | 0.07676 | 1563 | 28.34 | <.0001* |
| sex: female | 0.7867 | 0.2248 | 1558 | 3.50 | 0.0005* |
| sex: male | Ref |  |  |  |  |
| study_group: DP | -1.4332 | 1.0137 | 1562 | -1.41 | 0.1576 |
| study_group: ASAQ | -0.2494 | 1.0272 | 1564 | -0.24 | 0.8082 |
| study_group: standard care | Ref |  |  |  |  |
| ses: high | 0.2704 | 0.2240 | 1559 | 1.21 | 0.2277 |
| ses: low | Ref |  |  |  |  |
| age*DP | 0.1250 | 0.1072 | 1559 | 1.17 | 0.2438 |
| age*ASAQ | 0.002238 | 0.1082 | 1561 | 0.02 | 0.9835 |
| age*standard care | Ref |  |  |  |  |

DP= Dihydroartemisinin-Piperaquine, ASAQ=Artesunate-Amodiaquine, BMI=Body Mass Index, \*significant p-value, SES= Socio-Economic Status, Ref= Reference category.

**Table 7:**
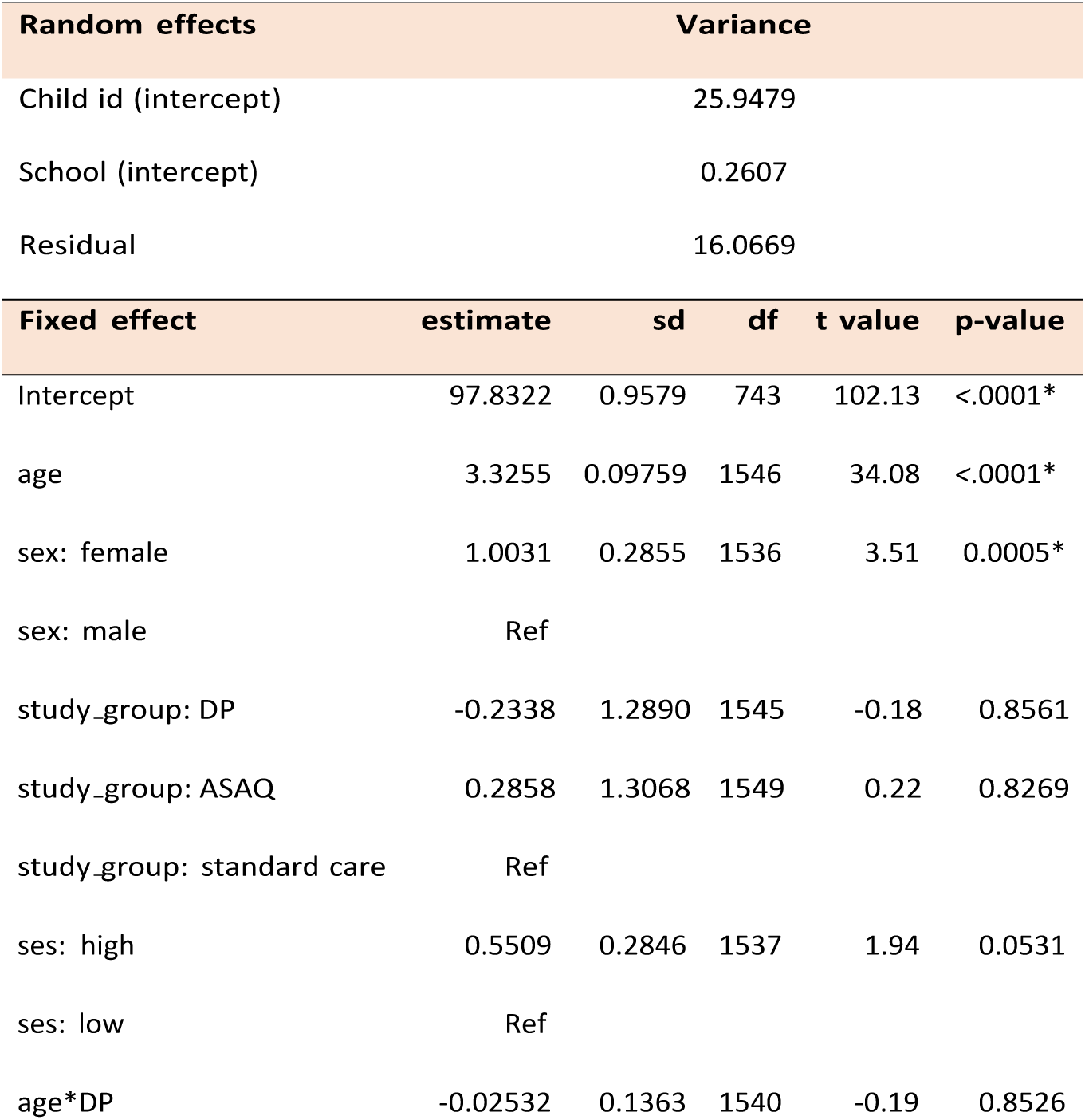
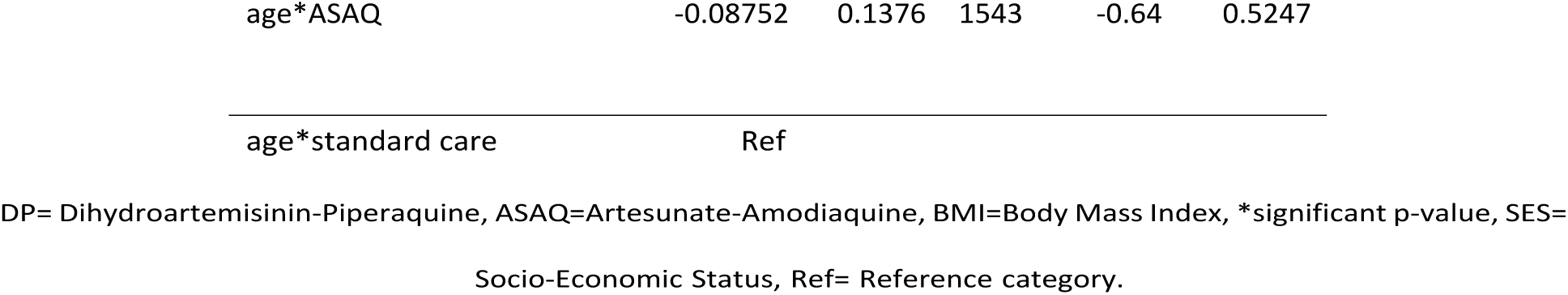
LMM results for height during trial.

**Table 8:**
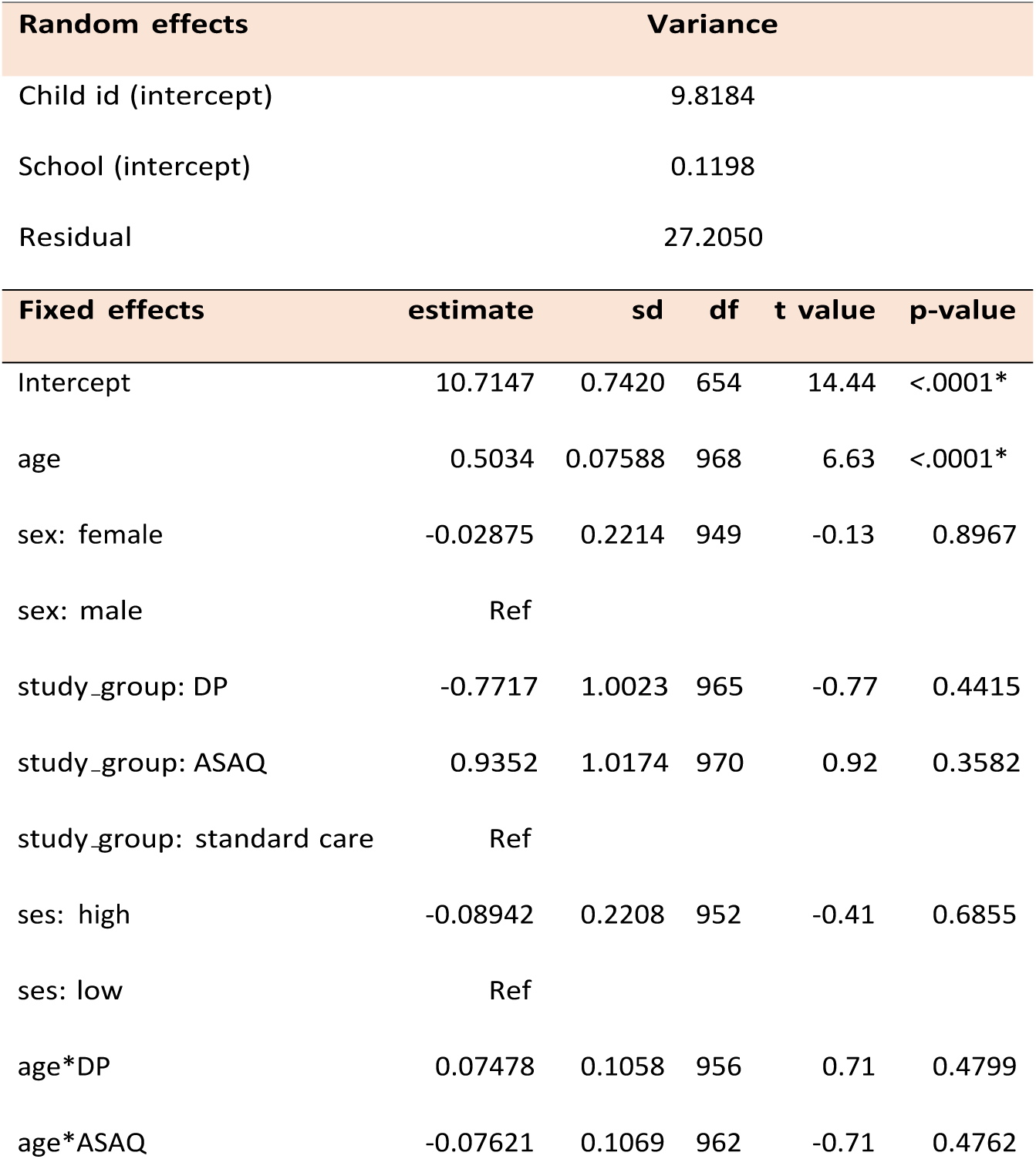
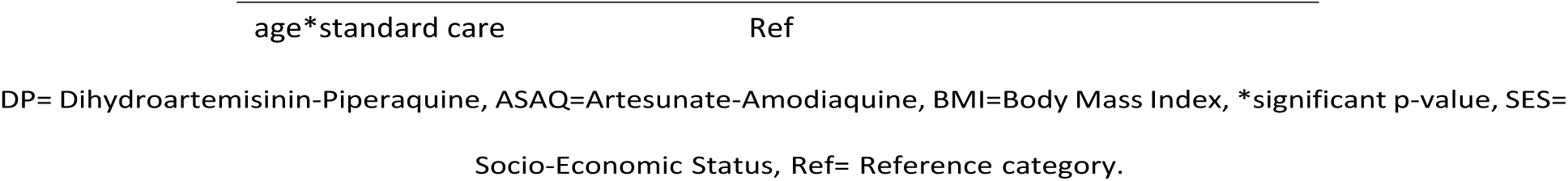
LMM results for BMI during trial.

When children were followed for 8 months after the intervention, there was no significant difference in weight, height, or BMI based on their former treatment allocation. The finding is intuitively reasonable because there was no actual treatment given to the participants. However, the effect of age on the growth parameters remained statistically significant even when treatments were stopped.

## 4 Discussion

Both malaria and malnutrition in children impose a burden of public health importance in LMIC in tropical regions. From the baseline data of our study, we found a malaria prevalence of 27%. 42% of children reported a history of malaria, and 13% used anti-malaria drugs in the past week. The prevalence of other parasitic infections, soil-transmitted helminths (STH), and schistosomiasis were 1% and 8%, respectively. For the nutritional status, 23% of 5-10 years old children were underweight (WAZ*<*-2), 21% of all children were stunted (HAZ*<*-2), and 28% were thin (ZBMI *<*-2). Most studies on malaria and malnutrition were conducted in children aged years and below, and one-third of children had at least one form of malnutrition in Tanzania [31]. In Nepal, 25.7% of adolescents were malnourished, and 21.8% were underweight [32]. A study conducted in Nigeria found malnutrition prevalence of 22% underweight, 37% stunting, and 6.9% wasted [33].

Among children diagnosed with malaria, 25% of 5-10 years old children were underweight, and 28% ofall children with malaria were stunted. While the malaria-malnutrition interaction has been observed in disease-endemic regions, it is difficult to establish which precedes the other [13]. In our study, we hypothesized that malaria infection (prolonged asymptomatic parasitemia in the malaria endemic region) poses a risk for malnutrition in children. However, most malaria-malnutrition studies investigated this phenomenon in children of age five years and below, with a perspective that malnutrition is the underlyingrisk for malaria since poor nutritional status leads to weak immunity [14, 34]. Malaria was significantly associated with stunting in our study. This finding is consistent with similar studies conducted in ≤5 years children that showed a significant association between malaria and being stunted. A cohort study that included a wide range of age (1 month to 14 years) in the Brazilian Amazon reported that multiple malaria episodes significantly negatively affected the linear growth of children of age 5 to 10 years [16]. Other observational studies from Sub-Saharan Africa in ≤5 years children reported a significant association [15, 35]. Stunting is a chronic condition that develops when an individual is exposed over a reasonably long time. Given that our data were cross-sectionally collected, the significant association between malaria infection diagnosed at one-time measurement is not biologically plausible. The finding rather reflects that children with recent malaria infection had previous recurrent infections since this study area is a stable malaria endemic region. During their life cycle, *Plasmodium falciparum* cause destruction of red blood cells leading to anemia. The subclinical hypoxic condition due to anemia inhibits insulin-growth factor-I, the hormone that plays a vital role in growth [36]. Such slowed growth experience predisposes children to long-term consequences of poor height gain depending on the level and duration of exposure.

We found an association between male sex and stunting as shown by higher odds and a p-value lower than5% significance level. The finding aligns with previous studies from children under five that the male sex was associated with at least one form of malnutrition [37, 38, 39]. We identified that children from low socio-economic status (SES) households had a statistically significant higher odds of being underweight. Prior studies also showed a significant association between low SES and underweight [31, 33]. Unlike underweight, stunting had no significant association with low SES [32, 33]. This insignificance implies that there could be factors that determine nutritional status in older children (school-age) that were not explored in the scope of this study, given that it was primarily designed for other purposes. Further nutritional studies need to investigate factors related to the life of a school-age child and adolescents as described by Scaglioni et al. in 2018 [40]; for example, distance from home to school, availability of food in a school setting, amount of food, meal frequency, and daily activities other than studies. Using a qualitative design that enrolls parents, teachers, and school children would be more informative.

Overall, an increase in weight and height in all participants was expected since age (a time factor) and sex are known to have a great influence on children’s growth. In our analysis using a linear mixed model, the effect estimates of DP and ASAQ on weight, height, and BMI were small and not statistically significant. Previous trials using anti-malarial drugs aimed at the effect of treatment in minimizing parasitemia, improvement of hemoglobin levels, and cognitive performance [4, 5, 19, 20]. The impact of treatment on nutritional status was reported rarely. Interventional studies that included infants and preschool children were conducted in Kenya [1997] and Senegal (2011), and they reported significant weight gain in treatment groups compared with the control group [41, 42]. The previous studies that reported significant weight and height gains in treatment groups included preschool children whose diet content and eating behaviors differ from the age group we studied. Energy requirements for children aged five and above are high, especially for children who live in low SES settings [43]. In that context, the difference in age-appropriate energy requirements, environment, and daily activities might have influenced the results.

Our study had several strengths; First, the use of both cross-sectional and longitudinal designs allowed assessment of risk factors and malaria-malnutrition burden. Knowing the prevalenceof malaria at baseline set the ground for the significance of doing the trial using anti-malarial drugs. The longitudinal design was useful for studying causal relationships. Secondly, to the best of our knowledge, this is among the few interventions to report the impact of intermittent preventive therapy for malaria on the nutritional status. This makes it a valuable knowledge source in addition to similar studies that included preschool children. The study does have some limitations. Firstly, we analyzed data that was primarily collected not for the purpose of nutritional assessment. For that reason, some known risk factors for malnutrition were not collected and, therefore, unavailable for analysis. In addition to known risks, it was important to assess other factors relevant to older children and adolescents, as it was shown in the Directed Acyclic Graphs (DAGs), Figure 1. Secondly, a wide range of available literature on the risks of malnutrition based on findings from under five years children who were extensively studied previously. Thislimits the comparability of information while discussing the findings of our study. Thirdly, the children in grades 6 and 7 were excluded from the study for practical reasons that they were anticipated to complete their school curriculum before the intervention study ended. The exclusion of older children limits the generalizability of results to the population of older children and adolescents. Another limitation was that, despite a significant association between malaria infection and stunting in our study, the conclusion on this causal effect is biologically incorrect because the parasitemia detected was measured only once at baseline. Restriction in height gain due to malaria infection can only be concluded given the exposure is stable for a considerably long time. Alternatively, this phenomenon could be investigated from a cohort design that takes into account the frequency of infections encountered by individual children. Lastly, we could not identify children who lived in the same household; therefore, clustering between households and variability between children from the same households could not be addressed in the analysis as a random effect.

## 5 Conclusion

The double health burden of malaria and malnutrition exists in almost equal proportion in the setting where we conducted the study. Factors known to be risks for malnutrition (low SES and Head of household education level) in under five years children were not significant in children aged ≥ 10 years. Further studies need to evaluate other factors relevant to the age group. For example, distance from home to school, availability of food in a school setting, amount of food, meal frequency, and daily activities other than studies. Using a qualitative design that enrolls parents, teachers, and school children would be more informative. The use of anti-malarial drugs for prevention purpose does not necessarily improve nutritional status. Combined with the previous findings suggestive of no established causal association between malaria infection and malnutrition, public health interventions need an integrated approach to address the two co-existing problems. Intuitively, malnutrition cannot be treated using anti-malarial drugs,and good nutritional status does not prevent malaria infection. The latter improves immunity to better fight infections, including malaria. Therefore, both malaria control and nutrition improvement programs should be implemented to achieve the intended health effect. The majority of the community members are peasants who produce food products for their families. Public health agencies should reinforce nutritional programs by collaborating with local communities to ensure food availability in schools and sustainable nutritional education to the local community members.

## Acknowledgement

I would like to express my sincere gratitude to the promotor of this study Prof. Jean-Pierre Van geertruyden, and Geofrey Makenga for their continued support throughout the development of this report. I would also like to thank Prof. Steven Abrams and Jackline Mbishi for their assistance in statistical analysis. In a special way, Iam grateful to the VLIROUS scholarship for providing financial support for this master’s program. These thanks are extended to the parties that helped in the acquisition of the data as mentioned in the other article [6]; the Muheza District officials; the District Medical Officer, district school health and NTD coordinators, schoolteachers, parents and guardians of children and the participants for their collaboration and support in the conduct of the study. Authors acknowledge stakeholders for school health program: The Ministry of Health, (MoH); the Ministry of Education, Science and Technology; the President’s Office-Regional Administration and Local Government (PO-RALG); the National Malaria Control Programme (NMCP) and the National Neglected Tropical Diseases (NTD) program for their cordial inputs with regard to study implementation and policy. The NIMR Director General and his staff for coordination of the study team and external stakeholders to the school health program. NIMR Tanga center staff for their cordial support during the preparation and implementation of the study. Global Health Institute staff (Universiteit Antwerpen, Belgium) for their administrative and logistical support.

## Data availability and sharing

The de-identified data will be available upon request. The approval for accessibility will be granted by the National Institute for Medical Research, Tanzania. This will involve signing data transfer agreement.

## Appendix

### Post trial

**Table 9:**
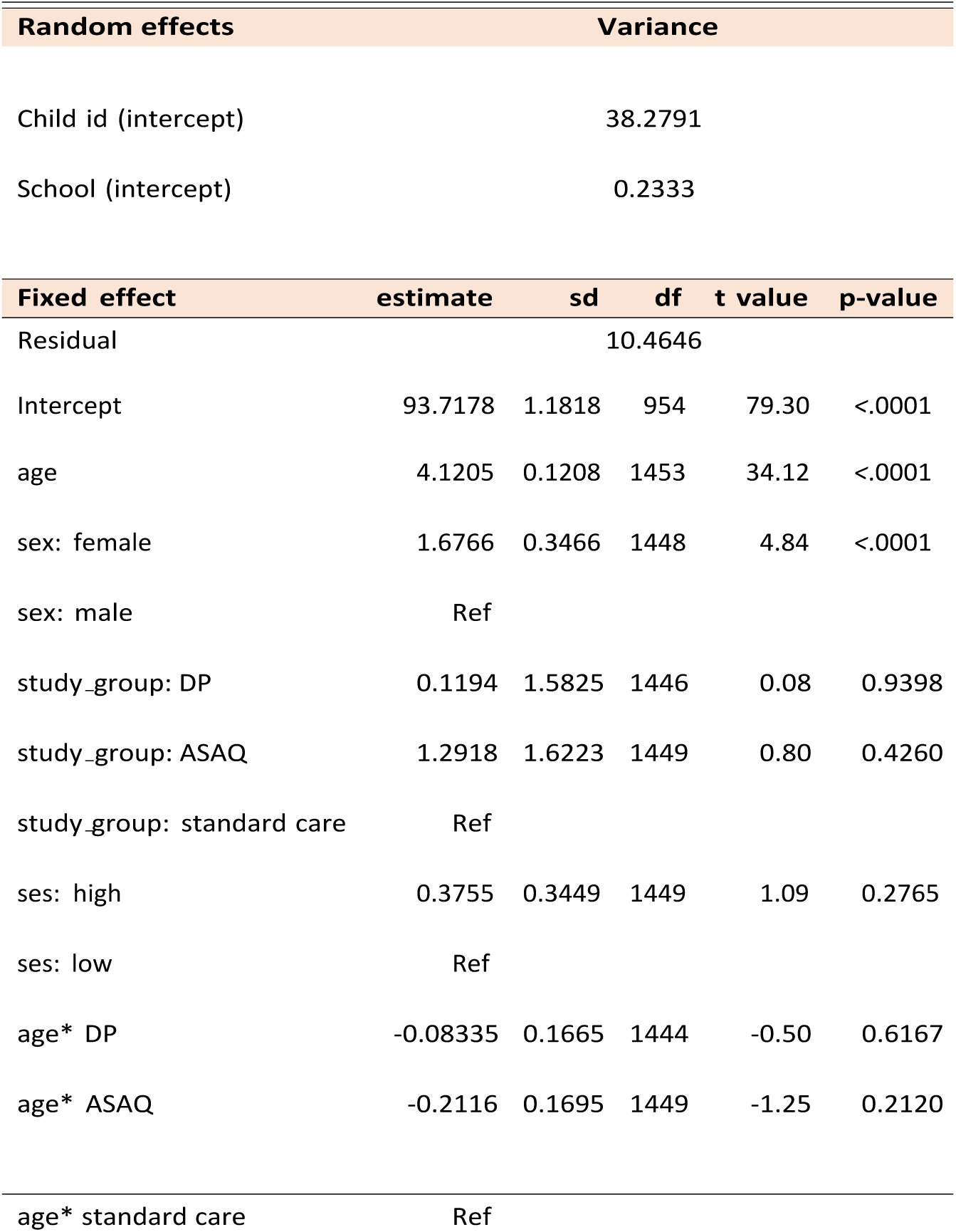
LMM results for height post trial.

**Table 10:**
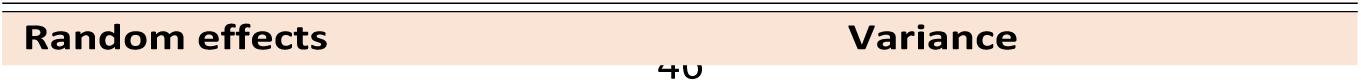
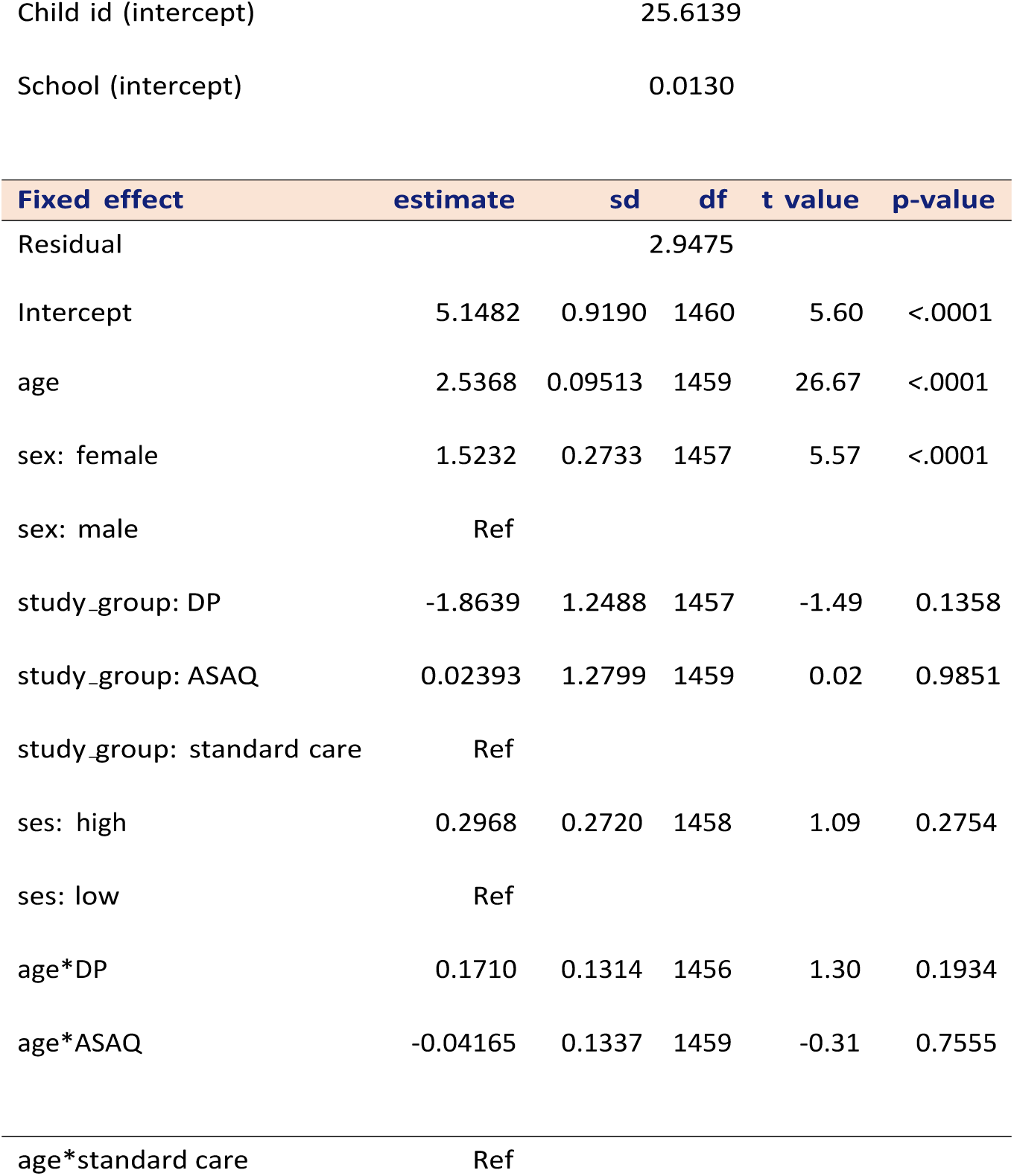
LMM for weight during post trial.

| Random effects | Variance |
| --- | --- |

|  |  |  |
| --- | --- | --- |
| 698 | Child id (intercept) | 25.6139 |
| 699 | School (intercept) | 0.0130 |

|  | Fixed effect | estimate | sd | df | t value | p-value |
| --- | --- | --- | --- | --- | --- | --- |
| 700 | Residual |  | 2.9475 |  |  |  |
| 701 | Intercept | 5.1482 | 0.9190 | 1460 | 5.60 | <.0001 |
| 702 | age | 2.5368 | 0.09513 | 1459 | 26.67 | <.0001 |
| 703 | sex: female | 1.5232 | 0.2733 | 1457 | 5.57 | <.0001 |
| 704 | sex: male | Ref |  |  |  |  |
| 705 | study_group: DP | -1.8639 | 1.2488 | 1457 | -1.49 | 0.1358 |
| 706 | study_group: ASAQ | 0.02393 | 1.2799 | 1459 | 0.02 | 0.9851 |
| 707 | study_group: standard care | Ref |  |  |  |  |
| 708 | ses: high | 0.2968 | 0.2720 | 1458 | 1.09 | 0.2754 |
| 709 | ses: low | Ref |  |  |  |  |
| 710 | age*DP | 0.1710 | 0.1314 | 1456 | 1.30 | 0.1934 |
| 711 | age*ASAQ | -0.04165 | 0.1337 | 1459 | -0.31 | 0.7555 |
| 712 | age*standard care | Ref |  |  |  |  |

**Table 11:** LMM results for bmi post trial.

|  | Random effects | Variance |
| --- | --- | --- |
| 714 | Child id (intercept) | 0.8612 |
|  | School (intercept) | 0.0211 |

| Fixed effect | estimate | sd | df | t value | p-value |
| --- | --- | --- | --- | --- | --- |
| Residual |  |  | 114.59 |  |  |
| Intercept | 12.0093 | 1.2986 | 1924 | 9.25 | <.0001 |
| age | 0.4086 | 0.1344 | 1925 | 3.04 | 0.0024 |
| sex: female | 0.7334 | 0.3854 | 1930 | 1.90 | 0.0572 |
| sex: male | Ref |  |  |  |  |
| study_group: DP | -1.0543 | 1.7561 | 1908 | -0.60 | 0.5483 |
| study_group: ASAQ | 1.2705 | 1.8052 | 1917 | 0.70 | 0.4817 |
| study_group: standard care | Ref |  |  |  |  |
| ses: high | 0.4647 | 0.3840 | 1936 | 1.21 | 0.2263 |
| ses: low | Ref |  |  |  |  |
| age*DP | 0.1101 | 0.1845 | 1901 | 0.60 | 0.5506 |
| age*ASAQ | -0.09210 | 0.1886 | 1916 | -0.49 | 0.6254 |
| age*standard care | Ref |  |  |  |  |

**Figure 5:**
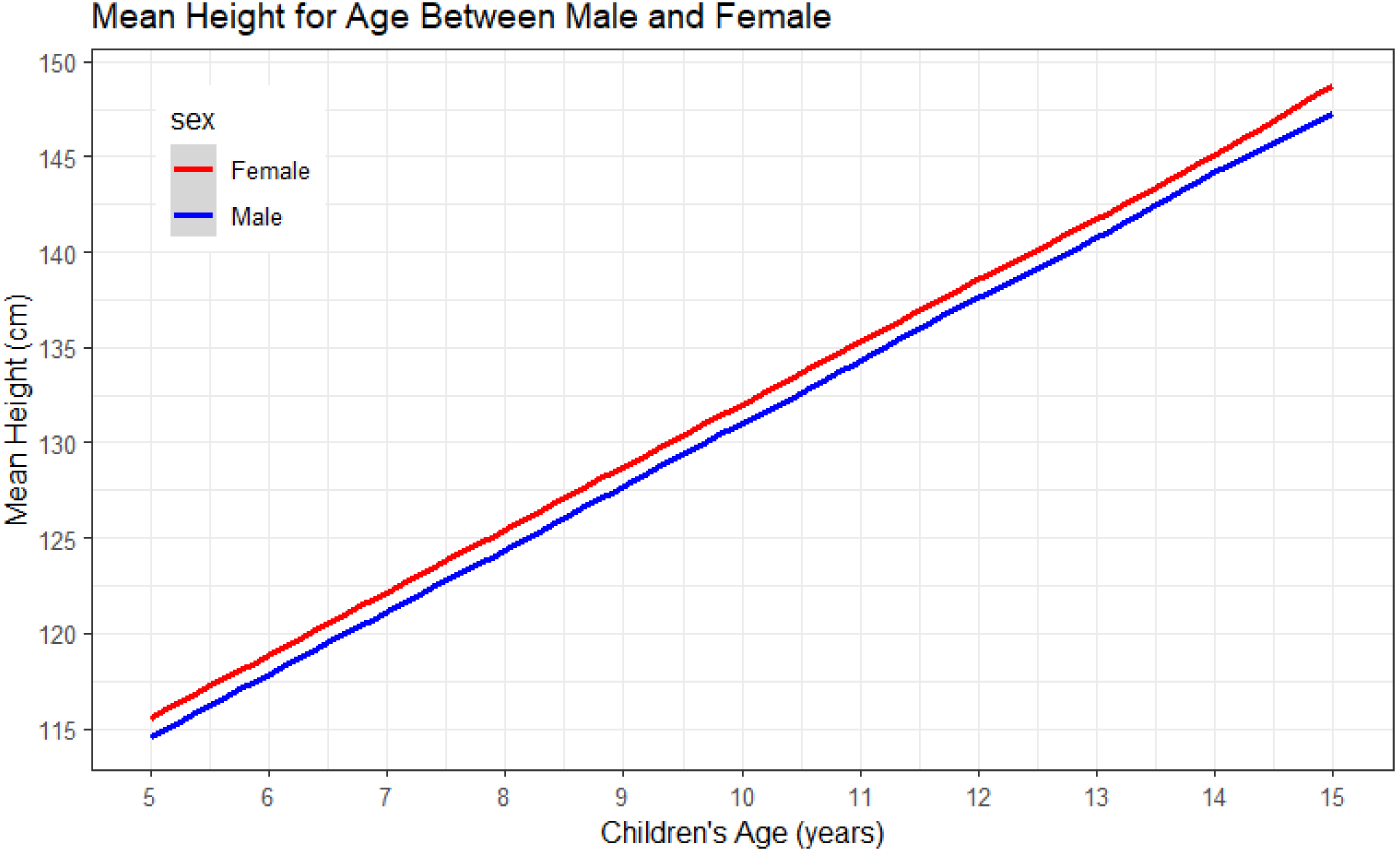
The trend of Height-for-age of school-age children grouped by sex

**Figure 6:**
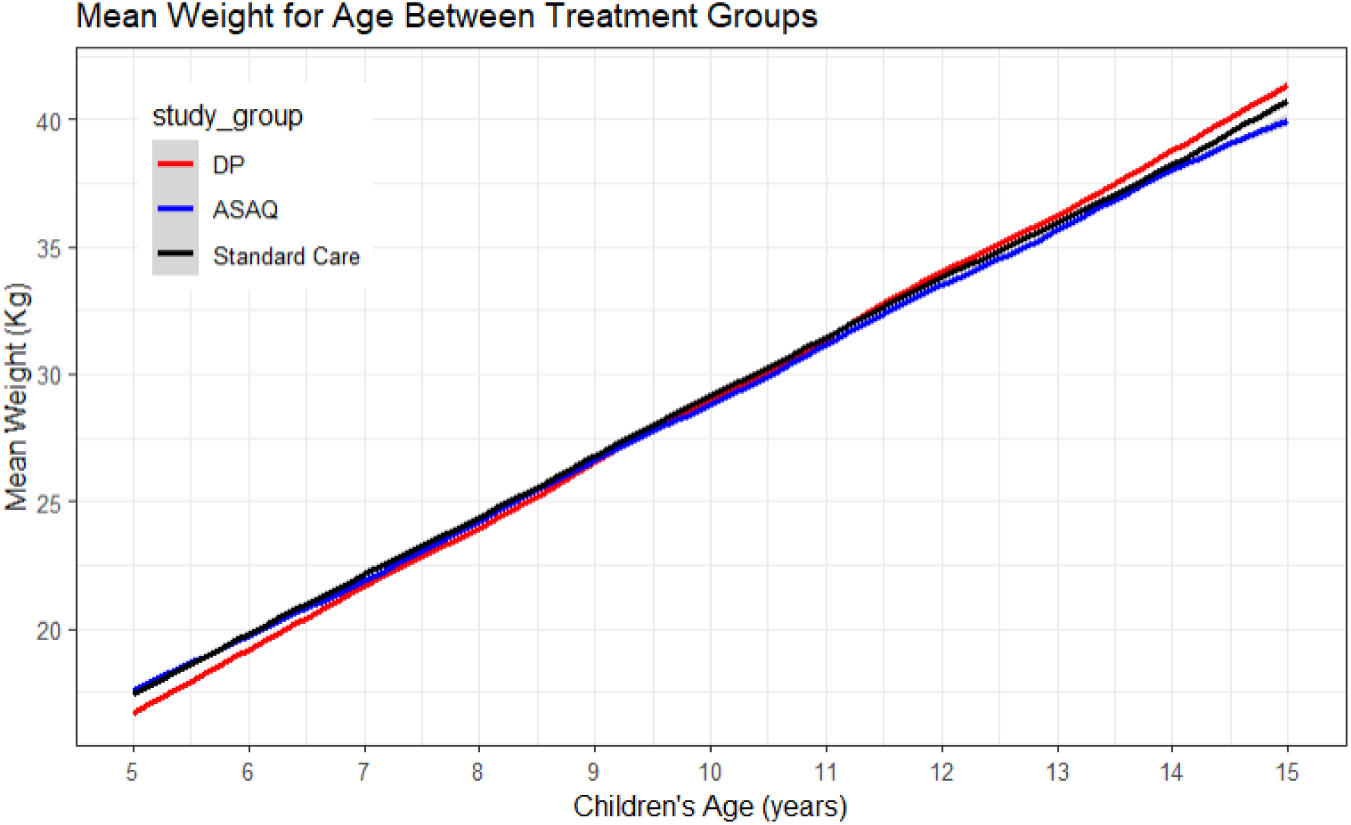
The trend of mean weight-for-age of school-age children grouped by treatment

**Figure 7:**
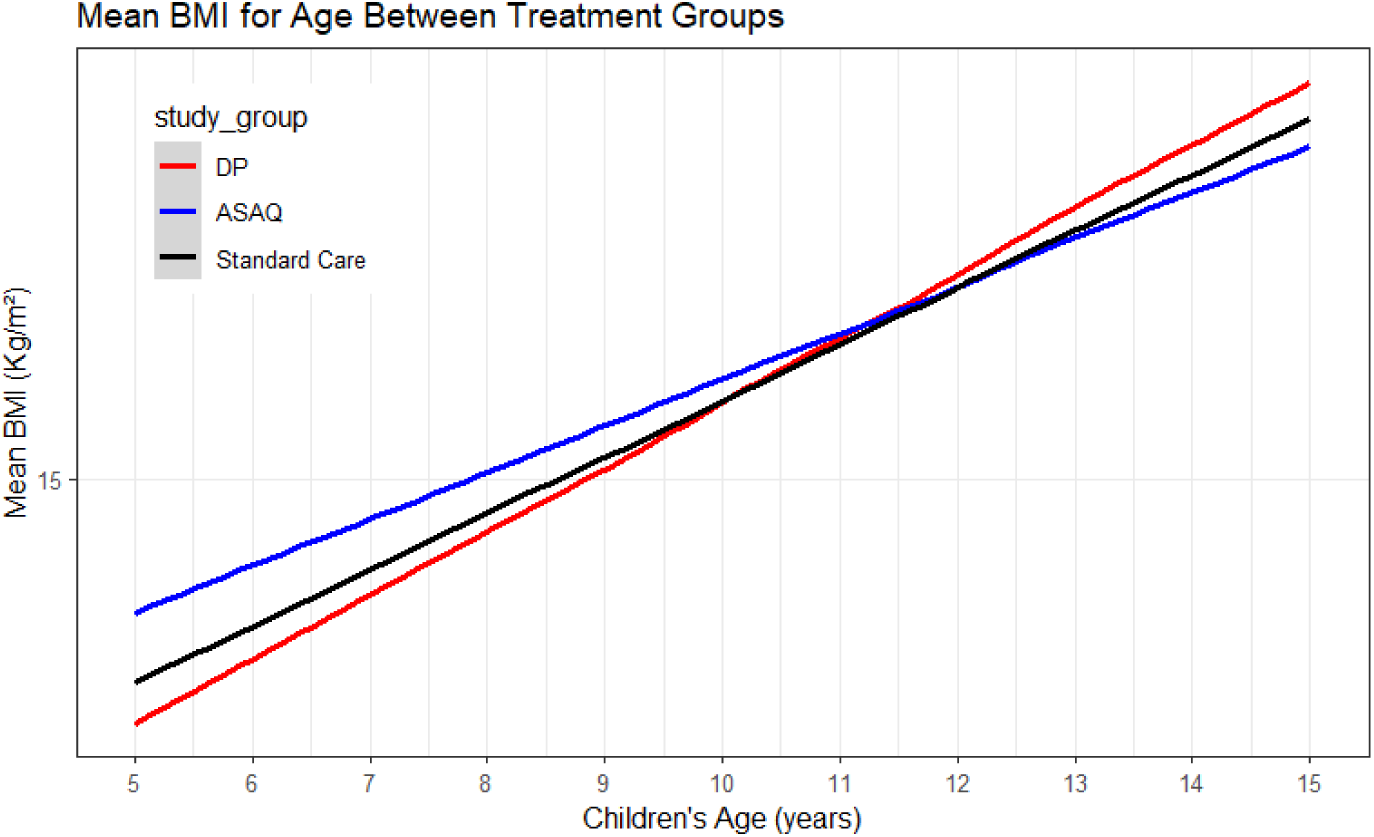
The trend of BMI-for-age of school-age children grouped by treatment

### Residual plots

**Figure 8:**
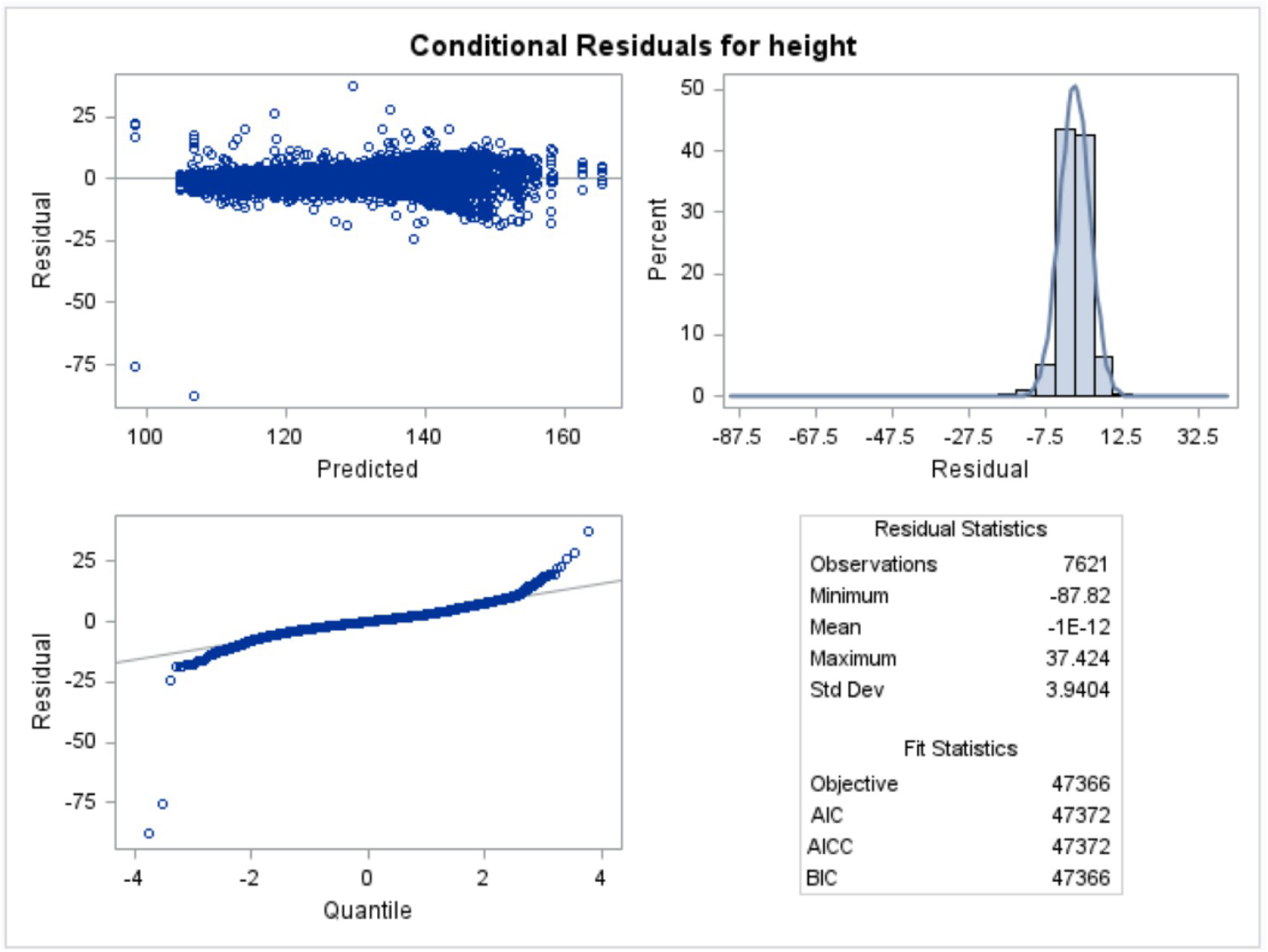
Residual plot for height

**Table 12:** AIC for LMM with different random effects.

| Models | child id | school | age | AIC |
| --- | --- | --- | --- | --- |
| 1 | intercept | intercept | slope | 48377.6 |
| 2 | intercept | intercept | - | 47372.3 |
| 3 | intercept | - | slope | 47423.7 |
| 4 | intercept | - | - | 48371.4 |

**Figure 9:**
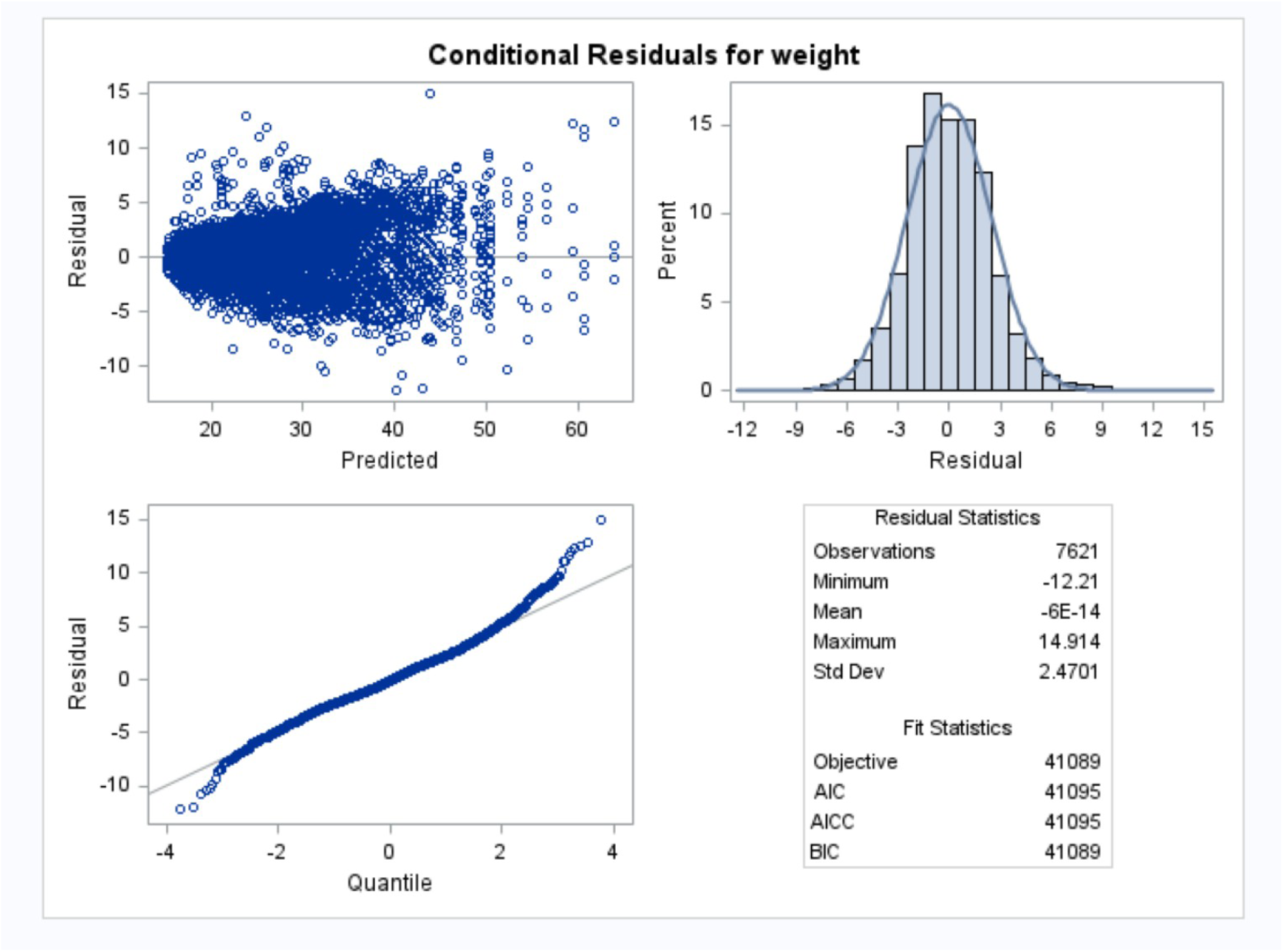
Residual plot for weight

**Table 13:** Comparing AIC values for models with and without quadratic term of age.

| Height | df | AIC |
| --- | --- | --- |
| Model1_age | 8 | 29839.3 |
| Model2_age<sup>2</sup> | 9 | 29837.5 |
| Weight |  |  |
| Model1_age | 8 | 24648.78 |
| Model2_age<sup>2</sup> | 9 | 24647.02 |
| BMI |  |  |
| Model1_age | 8 | 30711.92 |
| Model2_age<sup>2</sup> | 9 | 30711.50 |

